# High Resolution Multi-depth Quantification of the Retinal Nerve Fiber Layer

**DOI:** 10.64898/2026.05.22.26353127

**Authors:** Clémentine Callet, Maxime Bertrand, Kimberly Guzman, Pedro Mecê, Ethan A. Rossi, Kate Grieve

## Abstract

The retinal nerve fiber layer, composed of axon bundles converging toward the optic nerve, is a key biomarker for diagnosing and monitoring glaucoma and other neurodegenerative diseases. High-resolution en face imaging of individual nerve fiber bundles offers morphological information beyond what conventional optical coherence tomography provides, yet clinical integration remains limited by the lack of automated analysis tools and normative data. Here, we imaged 14 healthy volunteers using time-domain full-field optical coherence tomography and adaptive optics scanning laser ophthalmoscopy, and developed automated pipelines to quantify bundle width, trajectory, tortuosity, and orientation. Bundles were on average 25% wider at shallower retinal depths, width measurements were consistent across imaging modalities, and estimated axon count per bundle decreased significantly with age. Global trajectory analysis revealed systematic deviations of high resolution data from existing mathematical models, particularly in the temporal sector, leading us to propose two refined trajectory models. These normative results provide a foundation for high resolution biomarkers for use in investigations of retinal neurodegeneration.

---

Imaging and evaluation of the Retinal Nerve Fiber Layer (RNFL) is crucial in neuropathies because the retina is the only portion of the central nervous system accessible to direct optical imaging. Made up of Retinal Nerve Fiber Bundles (RNFB), each consisting of axons of retinal ganglion cells, the RNFL is responsible for transmission of visual information to the brain. Diseases such as multiple-sclerosis (*1*), Alzheimer’s (*2, 3*), Parkinson’s (*4*) and glaucoma (*5*) affect this layer of the retina. The diagnosis and follow up of neuropathic disease evolution can be assisted by imaging of this layer. Since the first optical coherence tomography (OCT) demonstration in 1991 by Huang et al. (*6*) and its commercial deployment in 1996, RNFL thickness measurements have become standard in clinical diagnosis, and the correlation between RNFL thickness and glaucoma, for example, is well established. Consequently since 2002, OCT RFNL thickness measurements constitute the main biomarker deployed worldwide to evaluate damage to the RNFL and progressive changes in disease (*7*). Adaptive optics can also contribute to ophthalmic diagnosis. Adaptive optics ophthalmoscopy (*8*) (AOO) provides cellular-scale retinal imaging beyond the resolution of conventional OCT. In particular, it allows the visualization of retinal ganglion cell (RGC) axon bundles and somas in vivo (*9, 10*), with the potential to resolve individual axons within sparse bundles. Owing to its larger FOV and high axial resolution, a high-resolution version of OCT named full-field optical coherence tomography (FFOCT) allowed bundle width to be assessed at different depths within the RNFL, offering good bundle separability in denser regions (*11, 12*).

The extraction of quantitative information from images of the RNFL is essential to improve our understanding of the mechanisms involved in disease progression and may also enable earlier diagnosis. If we narrow our focus to glaucoma, the second leading cause of blindness in developed countries, several potential biomarkers have been identified in recent years thanks to high-resolution imaging and advanced image-processing techniques. Firstly, the amount of light reflected from the RNFL, or reflectance intensity, appears to be linked to functional deterioration. This was measured using automated white-on-white perimetry with a Humphrey Field Analyser (HFA) (*13*). The HFA provides parametric visual field data expressed in decibels (dB), representing local differential light sensitivity. In this context, reflectance appeared to be associated with deep parametric defects, defined as locations with sensitivities below 15 dB (*14*). Furthermore, by combining this reflectance with RNFL thickness, RNFL Optical Texture Analysis (ROTA) has been developed, which allows for the detection of patterns of optical texture loss. This signature is associated with structural damage to RNFL axon bundles (*15*). To further characterize RNFB organization, a mathematical model of RNFB global trajectory was introduced by Jansonius and colleagues. It was based on fundus photography (*16*) and enabled a comprehensive description of the mean path and inter-trajectory variability of nerve fiber bundles. Subsequently, automated models were improved through the introduction of novel imaging techniques such as large field-of-view (FOV) polarization sensitive OCT (PS-OCT) (*17*), reducing variability associated with manual grading while eliminating time-consuming manual trace annotation. PS-OCT also demonstrated its relevance to assess damage in glaucomatous eyes by measuring retardance (*18*). Specifically, they found that a reduction in retinal retardance reflected not only thickness reduction but also microstructural axonal loss. Recent in-vivo imaging of mitochondrial function showed a relationship between mitochondrial dysfunction and glaucoma (*19*). The flavoprotein fluorescence (FPF) response was used to measure mitochondrial metabolic activity. Responses measured *in vivo* are reduced in glaucoma and glaucoma suspect eyes, indicating early retinal ganglion cell metabolic dysfunction associated with glaucoma.

Beyond these exploratory studies, another potential RNFL biomarker accessible with high resolution imaging is the diameter or width of the individual bundles and their organization. For routine measurement of RNFL structure, conventional OCT is currently the gold standard even though it cannot achieve cellular resolution. Indeed, a substantial portion of RGCs must be lost (10–40%) before a statistically significant thinning can be detected (*20, 21*) with OCT, limiting it to the detection of global, significant changes only (*22, 23*). Moreover, RNFL thickness varies among individuals depending on age, sex, refractive error and ethnicity (*24–28*), which further hinders early diagnosis. On the other hand, high-resolution imaging techniques can detect subtle structural changes, but currently lack tools for quantification. As these high-resolution imaging techniques arrive in the clinical setting, we require new tools to analyse them.

In this study we explore a number of structural RNFL parameters in high resolution and standard clinical images. We aim to obtain precise structural information on the RNFL, specifically the RNFBs, to establish baseline measurements of healthy individuals to serve as a reference for comparative analysis to individuals with neuropathies. We also aimed to better understand RNFL fiber bundle structure in depth. We used a FFOCT (*12,29*) system, two AOSLOs (one research-grade and one commercial, with similar specifications, used on different parts of the study depending on availability), and a cSLO-SDOCT to image the RNFL in healthy volunteers across a range of ages. We developed tools that are automatic and semi-automatic to quantify high resolution images. We propose a model to quantify bundle orientation in normal retina which could then signal regions of abnormality in disease.

## Results

After coarse registration, high-contrast images of RNFL axon bundles were obtained on all participants. At 4°N, 17 research-grade AOSLO images and 99 FFOCT images from 11 healthy volunteers were quantified effectively. 396 Mona IV AOSLO images from 9 participants were quantified from 12°N to 10° T and 6° Superior to −3° inferior. RNFL thickness heatmaps were generated from Spectralis OCT data on 7 of the participants.

### RNFB width

Automatic segmentation allowed us to follow the bundle paths and quantify their width. 2046 bundle segments were successfully measured (443 with research AOSLO, 1603 with FFOCT). The 8 μm optical sectioning ability of FFOCT enabled quantification of bundle width through a 50 μm depth range which increased the number of bundles quantified compared to AOSLO, which with its optical sectioning of 40μm includes the whole RNFL thickness in a single en face plane. Note that depth sectioning with FFOCT is only feasible in regions where the RNFL is thick enough to support such an axial extent. We defined the shallow layer as the first layer after the ILM in which RNFBs could be quantified, and the deepest layer as the last layer in which RNFBs could be quantified before the RNFB signal disappeared. All intermediate layers quantified in this study are indexed and represented in a gradient color from green (shallow) to blue (deeper) (cf Figure 1). They correspond to the remaining RNFL, divided into segments of equal thickness. We compared the averaged bundle width measured on AOSLO and FFOCT in the exact same ROI on participant #4 (Figure 1.A.). First, the automatic measure on the averaged bundle width on the AOSLO image gave 30 μm which is less than the shallow layer measurement with FFOCT. When going deeper, the average width decreased, until the deepest layer (FFOCT 4) where only the wider RNFB remained visible on the left, and on which we can see vessels appearing already on the right. Analysis of the full cohort revealed that nerve fiber bundles were 25% wider, on average, at the inner retinal layers compared to the deeper layers for the same eccentricity (Figure 1.B.). In detail, Δ*width*_*deep*−*shallow*_ = −9.95*µm* ± 6.04*µm* (for 29 ROI) which corresponds to a width decrease of 25% with increasing depth (Δ*width*_*deep*−*shallow*_ = −25.16% ± 13.01%.)

**Figure 1.**
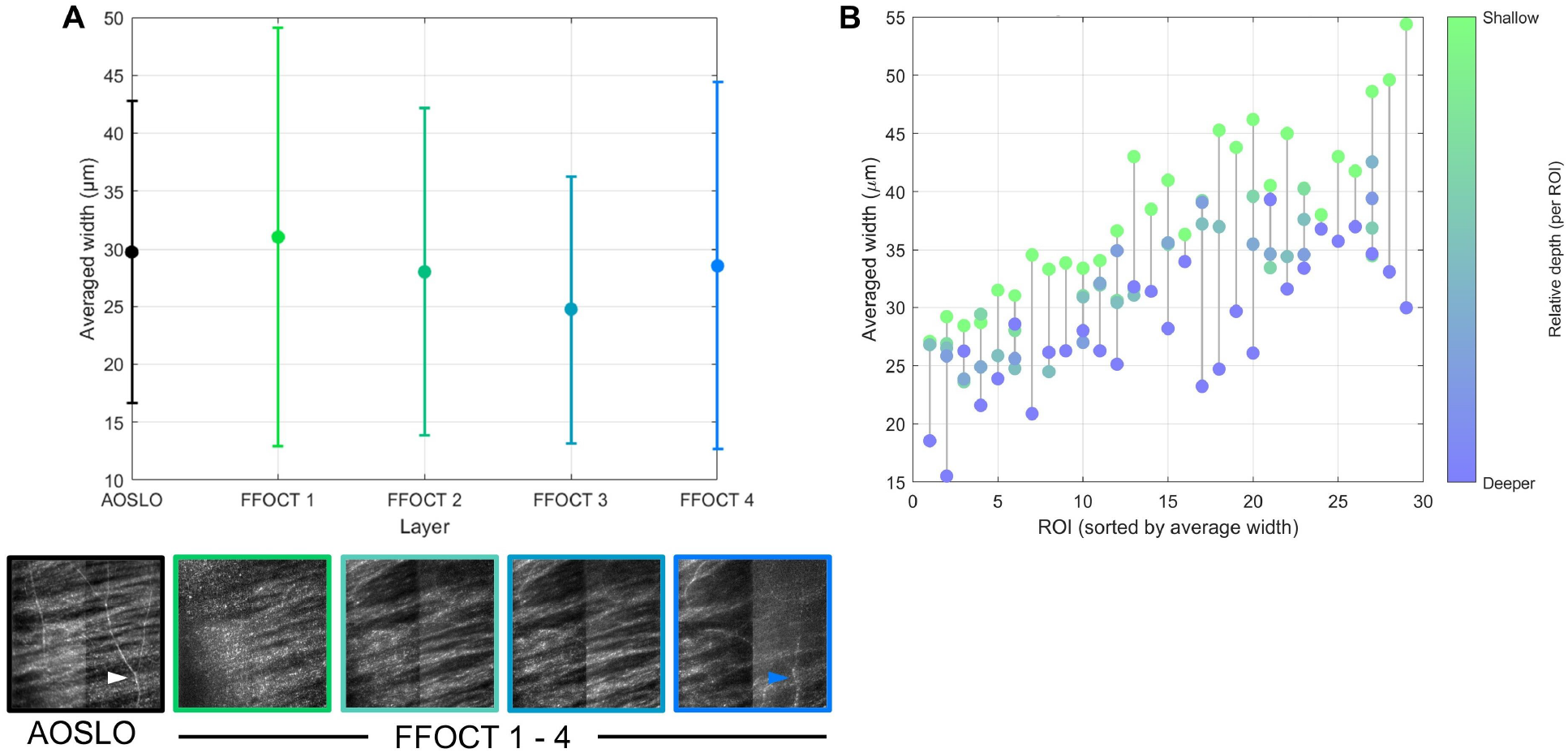
Deeper bundles are thinner. The figure illustrates RNFB width quantification using research AOSLO and FFOCT at different depths. (**A**) Same region of interest (1.5° × 1.5°) in participant #4 imaged with AOSLO (single depth) and FFOCT (four depths from superficial to deep). (**B**) Collection of 29 FFOCT ROIs acquired at different depths across 11 participants. “Shallow” corresponds to the first layer below the internal limiting membrane (ILM) where RNFBs could be quantified, whereas “deep” corresponds to the last layer where RNFBs remained detectable before signal disappearance. Intermediate colours represent quantifiable layers between these two extremes.

If we now examine the average bundle width per depth, sorted by age (Figure 2.A), there appears to be an age-related effect: the average bundle width declines more markedly at relative depths 4 and 5 with increasing age, whereas it remains relatively stable at relative depths 1 and 2.

**Figure 2.**
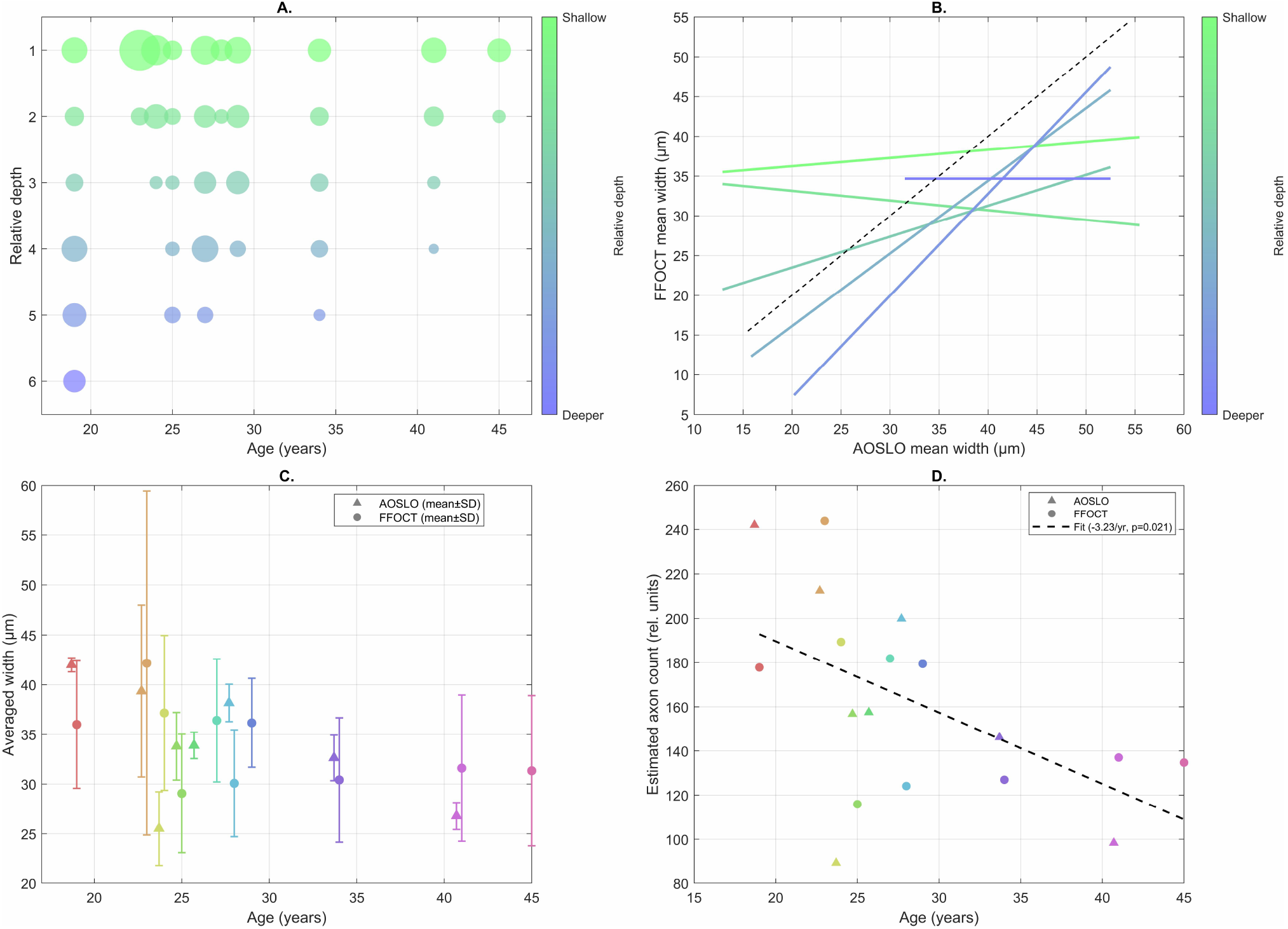
Bundles in older eyes are thinner. The figure compares RNFB width estimates derived from FFOCT and research-grade AOSLO imaging. (**A**) Effect of imaging depth on RNFB width measured using FFOCT data only; circle size represents the average bundle width. (**B**) Comparison of RNFB width measured at different depths using FFOCT and corresponding measurements from research AOSLO. (**C**) Mean RNFB width per participant, averaged across all ROIs located around 4° nasal, measured using research AOSLO (triangles) and FFOCT (points). (**D**) Estimated axon count per bundle, derived from average RNFB width measured with AOSLO (triangles) and FFOCT (points). A linear fit across both datasets indicates a decrease of 3.23 axons per bundle per year, assuming an axon diameter of 2.7 μm (*30*).

Overall, bundle widths measured by both FFOCT and AOSLO modalities were of comparable magnitude, ranging from approximately 15 to 55 μm, although strict agreement with the identity line was observed in only a minority of bundles (Figure 2.B). Superficial bundles (green) tended to show slightly higher FFOCT values relative to their AOSLO counterparts, lying at or above the diagonal. In contrast, deeper bundles (blue) tended to display larger AOSLO values than FFOCT. To compare AOSLO and FFOCT averaged width results we chose the layer acquired at depth offset 5, corresponding to a relative depth of 10 *µ*m below the ILM. We processed 46 ROIs located around 4°N and quantified the average bundle width as a function of age (Figure 2.C). A global decrease in width with age was observed. The average width measured by both modalities was very similar. AOSLO *averaged_width*_*AOSLO*_ = 33.93*µm* ± 6.08*µm* and FFOCT *averaged_width*_*FFOCT*_ = 32.92*µm* ± 7.07*µm*. When comparing values measured per individual, with FFOCT and AOSLO across ROIs, we found Δwidth_|FFOCT−AOSLO|_ = 5.75 *µ*m ± 3.25 *µ*m, on average for our cohort, with a maximum difference of 11.6 *µ*m (participant #3 shown in yellow in Figure 2.C) and a minimum of 2.2 *µ*m (participant #6 shown in purple Figure 2.C). These differences arise from the use of different imaging instruments with different axial resolutions. We then aimed to quantify the age-related axon loss observed in our cohort. To do so, we assumed that axons are cylindrical with a diameter of 2.7 μm (*30*), and that RNFBs are cylindrical with an average diameter equal to the average width we measured. Based on these two assumptions, we estimated the number of axons in a single bundle by dividing the bundle cross-sectional area by the axon cross-sectional area, both modeled as circular sections. A linear fit revealed a significant decline in axonal number over time (p = 0.021), corresponding to an estimated loss of 3.23 axons per bundle per year (Figure.2.D). We lastly examined the relationship between averaged RNFL thickness measured by the Spectralis and averaged bundle width measured with the high resolution modalities. A linear regression analysis revealed no significant association, i.e. layer thinning is not equivalent to bundle thinning. Taken together, these results indicate that while FFOCT and AOSLO capture broadly comparable bundle width trends across the population, including a decrease in width with age, differences remain at shallow depths, reflecting differences in contrast and axial resolution inherent to each modality.

### Local and global RNFB trajectory and tortuosity

Our automatic segmentation measured both local and global trajectories by extracting nerve fiber bundle paths. The local trajectory, measured on research AOSLO (Figure 3) and FFOCT corresponds to the locally computed orientation derived from the segmentation, whereas the global trajectory (Figure 4) corresponds to the resulting traces obtained from the orientation map computed from wide–FOV segmented Mona IV AOSLO images combined with a mathematical model. Assessing local trajectory allows for quantification of subtle changes. At 4° nasal, for ROIs from FFOCT, inter-participant variability in tortuosity was low, with values between 1.03 and 1.06 (mean 1.03 ± 0.003 SD), and RNFB trajectories were close to 0°, spanning from −1.96° to 0.77° (mean 1.05° ± 0.01° SD). Similarly, AOSLO ROI measurements acquired at 4.5° nasal and 1° inferior showed low tortuosity values, between 1.03 and 1.06 (mean 1.04 ± 0.01 SD), with trajectories also close to 0°, extending from −2.7° to 4.42° (mean 0.27° ± 2.7° SD). When comparing FFOCT and AOSLO at matched retinal locations, tortuosity values were comparable while RNFB trajectories showed slightly larger variations. Overall, FFOCT and AOSLO showed good agreement; therefore, AOSLO was used for the remainder of the trajectory measurements in this study. Figure 3 is an example of the local trajectory algorithm result. It is an AOSLO montage of 30 images acquired on participant #5. From 14°N to 6°N, we ran the full pipeline and computed the local orientation/trajectory in degrees. The zero corresponds to the result of our disc-fovea angle computation described in the methods. We also looked for a significant relationship between width and orientation, and width and tortuosity, by performing a linear regression analysis and using linear mixed-effects models. Point-wise morphological analysis was performed along the skeletons of individual RNFL bundles across 11 participants, yielding *N* = 112,604 valid measurement points. Global linear regression revealed statistically significant but negligible associations between local bundle width and both orientation (*R*^2^ = 0.0007, *p* < 0.001, slope = +0.083 *µ*m/deg) and tortuosity (*R*^2^ < 0.0001, *p* = 0.025, slope = +1.56 *µ*m), with Spearman correlations of *ρ* = 0.029 and *ρ* = 0.009 respectively, confirming the absence of any clinically meaningful linear relationship. Per-participant analysis showed no consistent directional effect, with slopes alternating in sign across individuals and no participant exceeding *R*^2^ = 0.012.

**Figure 3.**
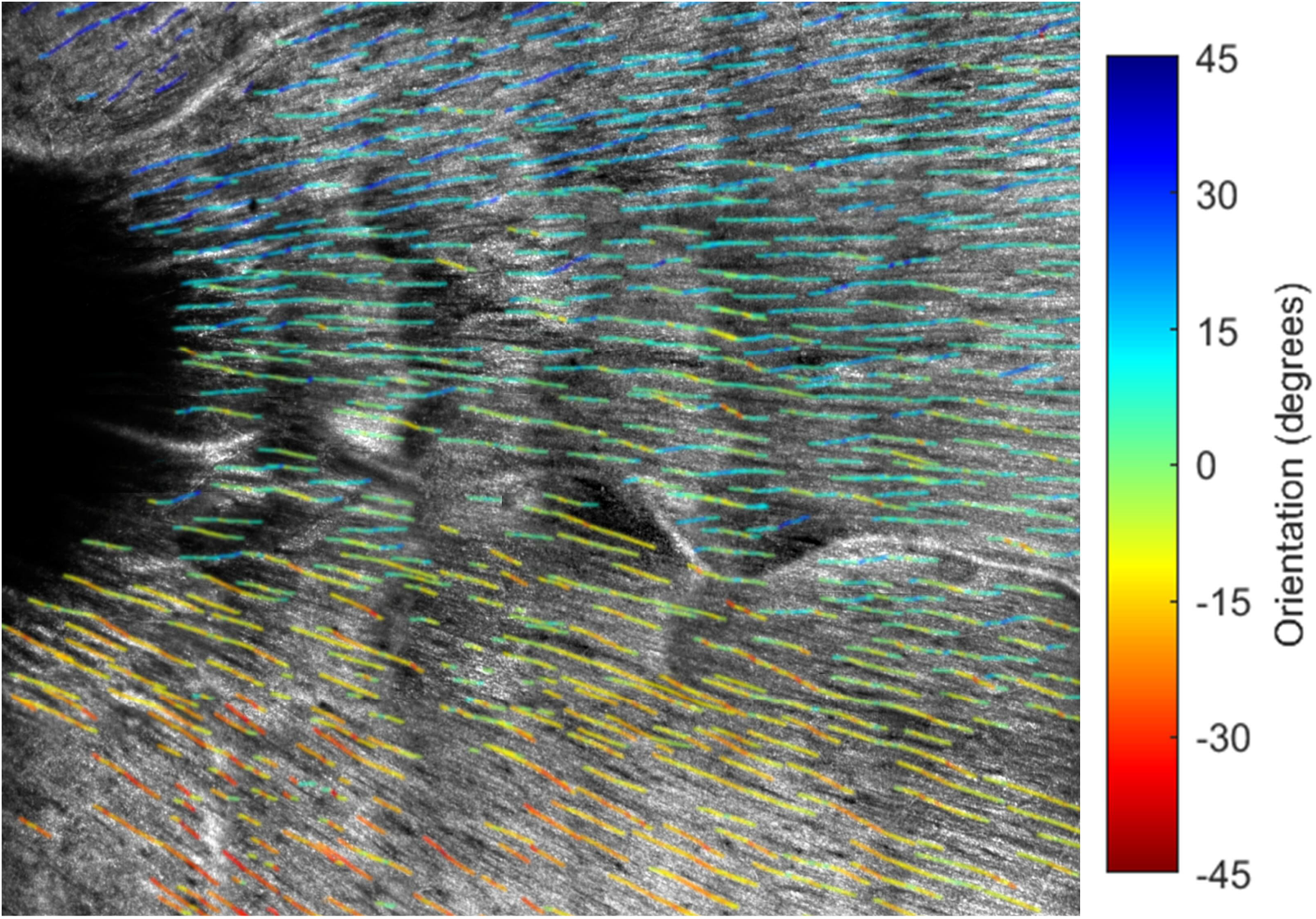
Local orientation of RNFB. Retinal Nerve Fiber Bundle trajectories for participant #5, segmented and reconstructed from research-grade AOSLO images acquired in Pittsburgh (cf Methods section Local orientation and Figure 8)

**Figure 4.**
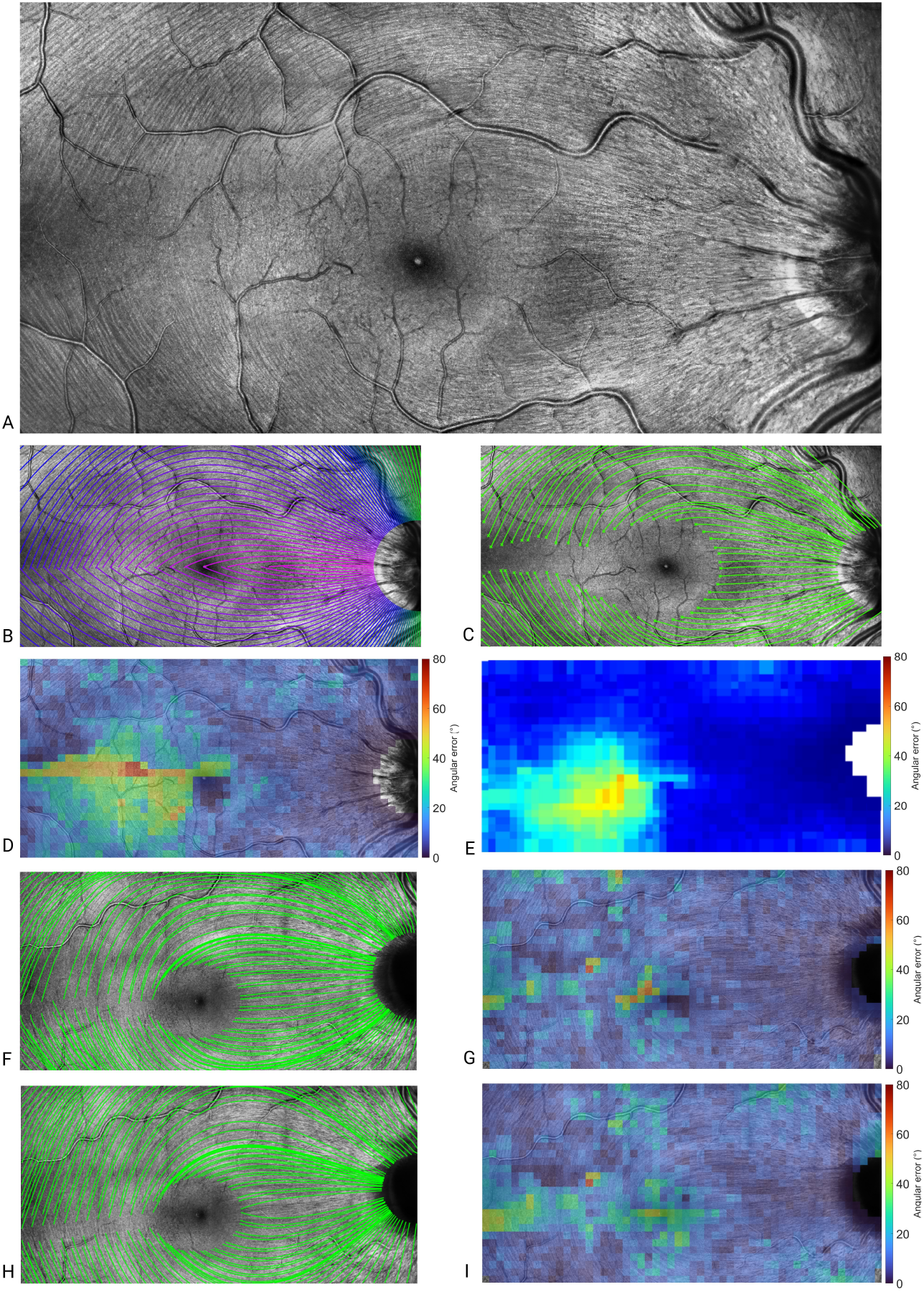
Comparison of RNFB trajectory models on a wide field fundus montage. The montage (14°x 26°) is overlaid with different RNFB trajectory estimations and their corresponding angular differences. (**A**) Original montage. (**B**) Overlay with the mathematical model described by Jansonius et al. (*16*). (**C**) Overlay with trajectories derived from our proposed method based on Schwarzhans et al. (*17*), initialized from manually selected starting points. (**D**) Colour map of the angular differences between (B) and (C). (**E**) Heat map of the mean angular difference across all participants between the Jansonius model and our segmentation. (**F**) Result of the proposed Bézier-based RNFB model applied to participant #14, not included in the training dataset. (**G**) Colour map of the angular differences between (F) and (C). (**H**) Result of the fine-tuned Jansonius model applied to participant #14 using the same initialization points. (**I**) Colour map of the angular differences between (H) and (C).

Linear mixed-effects models accounting for the nested structure of intrabundle intraparticipant measurement points confirmed the absence of a significant fixed effect of orientation on local bundle width (slope = −0.006 *µ*m/deg, *p* = 0.602). A small but significant effect of tortuosity was retained (slope = +3.83 *µ*m, *p* < 0.001), however the marginal *R*^2^ of 0.034 indicated that tortuosity explained a negligible fraction of inter-bundle width variance. Variance decomposition revealed that inter-bundle variability (variance ≈ 5.0) accounted for approximately six times more variance than inter-participant variability (variance ≈ 0.8), meaning that bundle identity is the primary determinant of local width. Finally, analysis of the 90^th^ percentile envelope revealed a consistent boundary effect for tortuosity: the maximum achievable bundle width decreased markedly for tortuosity values exceeding approximately 1.25, from 45 *µ*m to 25 *µ*m, indicating that tortuous bundles are thinner. This pattern was reproduced across all participants.

To evaluate global trajectory, orientation maps (OM) of RNFB traces were computed for 9 participants. To cover the fundus efficiently for tracing of global trajectories, we chose lower resolution 5° FOV tiles on MONA IV AOSLO to stitch a wide field fundus image (Figure 4.A). For each montage, a personalized OM was created; using selected points on the image, a path could be traced for each bundle. The result of the difference in orientation between Jansonius orientation map *OM*_*M*_ (Figure 4.B) and our computed orientation map *OM*_*S*_ (Figure 4.C) is pictured in Figure 4.D. The mean inter-participant angular error for each zone is presented in Table 1 and Figure 4.E. The last participant we imaged (participant #14) was used to evaluate the final model we obtained (cf Figure.4.F-G).

**Table 1.** Angular deviation error between Jansonius et al. map and the segmented map. Retinal nerve fiber bundle (RNFB) orientation maps were compared to the Jansonius model (*16*) by retinal sector (fundus coordinates, right eye). Values are group medians, interquartile ranges (IQR), and 90th percentiles (P90) across nine participants. Polar angle convention: 0° = nasal, 90° = superior, 180° = temporal, 270° = inferior. Sectors sharing a superscript letter do not differ significantly (Kruskal–Wallis *p* < 0.001, post-hoc Bonferroni correction).

| Sector | Angle range | Median error ($^\circ$) | IQR ($^\circ$) | P90 ($^\circ$) |
| --- | --- | --- | --- | --- |
| Temporal | $135^\circ$–$225^\circ$ | $25.2^a$ | 4.4 | 34.0 |
| Inferotemporal | $225^\circ$–$270^\circ$ | $28.5^a$ | 10.0 | 38.5 |
| Superonasal | $0^\circ$–$90^\circ$ | $6.0^b$ | 1.7 | 15.1 |
| Inferonasal | $270^\circ$–$360^\circ$ | $6.3^b$ | 1.2 | 13.2 |
| Superotemporal | $90^\circ$–$135^\circ$ | $10.7^b$ | 2.5 | 21.3 |

Across the nine participants, the median angular deviation between the measured orientation maps and the Jansonius et al. (*16*) model was 9.4° (IQR: 9.4–10.1°), indicating a moderate but consistent divergence at the population level. A significant asymmetry was observed between hemifields: the inferior hemifield exhibited substantially larger errors (median: 11.9°) than the superior hemifield (median: 8.8°), a difference that was statistically significant (Wilcoxon signedrank test, p = 0.0039). All nine participants showed the same direction of asymmetry. Sector-level analysis revealed a striking spatial pattern of deviation from the Jansonius model (Table 1, Kruskal-Wallis *p* < 0.0001). The largest errors were concentrated in the Temporal sector (median 16.5, IQR 3.6) and the Inferotemporal sector (16.3, IQR 4.2), both of which were significantly higher than all three remaining sectors (Bonferroni post-hoc, all *p* < 0.04). By contrast, the Superotemporal (5.1°), Inferonasal (5.6°), and Superonasal (6.3°) sectors showed consistently low errors with no significant differences among them. The spatial distribution of errors in polar coordinates (Figure 4.D) revealed that the largest deviations were concentrated around the horizontal meridian at eccentricities between 8°T and 12°T. In contrast, regions corresponding to the arcuate fiber bundles showed comparatively lower errors at mid-eccentricities, though the inferior arcuate region consistently exhibited higher errors than its superior counterpart, in line with the hemifield asymmetry reported above. This spatial pattern (cf Figure 4.D) was conserved across all participants. Our Jansonius model fitting provided a new equation for Jansonius trajectories defined as 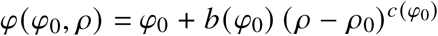 with 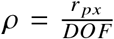, where *r* _*px*_ is the radial distance in pixels from the ONH center and *DOF* is the distance between the ONH center and the fovea, and with 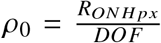, where *R*_*ONHpx*_ is the measured ONH radius in pixels. *c*(*φ*_0_) and *b*(*φ*_0_) were defined as:

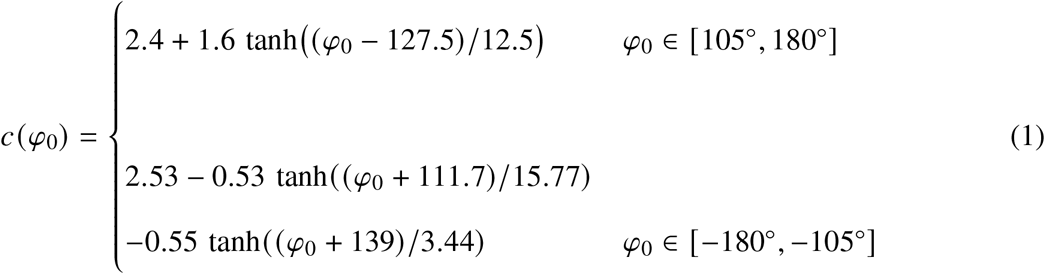

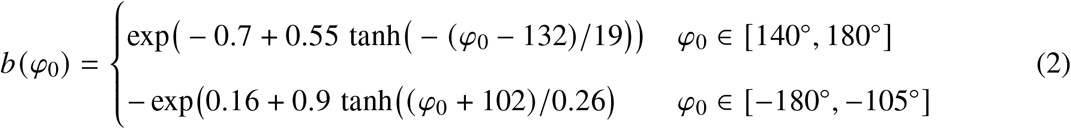

Finally, we compared the original Jansonius model, our refined parametric Jansonius model, and the Bézier-based approach (see Methods) on participant #14, who was not included in the dataset used to build the model (Figure 4.F-I). The original Jansonius model exhibited substantial errors, particularly in the temporal retina (22.92°) and inferior regions (19.15°), with a strong deviation in central areas (19.15° vs 15.14° in the periphery). In some localized regions, errors reached up to 67°. The refined Jansonius model significantly improved the fit, especially in the periphery, where the error decreased from 15.14° to 10.56° (30% reduction). The temporal quadrant showed the most notable improvement, dropping from 22.92° to 13.04° (43% reduction). Errors also became more homogeneous across quadrants (10°–13° range), although central errors remained relatively high (18.17°). The Bézier model provided the best overall performance. It further reduced the central error to 14.48° and the peripheral error to 8.92°, corresponding to a 25% improvement over the refined Jansonius model and more than 40–50% compared to the original model at the raphe. The nasal quadrant reached the lowest error (6.67°), while temporal errors remained comparable to the refined Jansonius model (13°). Peripheral nasal regions showed the strongest gain (down to 5.25°). Overall, while the refined Jansonius model corrects a large portion of the systematic bias of the original formulation, particularly in the temporal retina, the Bézier approach consistently achieves the lowest errors and provides the most accurate description of RNFB trajectories across the full field of view. We suggest using the refined Jansonius model when an instantaneous overlay of RNFL trajectories on an image is needed for overall comparison or RNFB mapping. For more precise quantitative analyses, we recommend using the Bézier model to compare the patient OM with the Bézier OM. In both approaches, only three manual clicks to identify the fovea, ONH center, and ONH radius are required for the OM generation.

Other parameters analysed on the wide FOV AOSLO montages were the raphe and disc angles. The raphe angle relative to the horizontal was −3.3° ± 3.4° (range: −8.7° to +1.2°), consistent with Huang et al. (*31*) who reported −1.67°± 4.8°. The raphe-fovea-disc angle was 170.45°± 3.5° (range: 163.3° to 178°) which is also consistent with Huang et al. who reported 170.3°± 3.6°.

## Discussion

In this study, we demonstrate that high resolution imaging enables reliable quantification of the individual retinal ganglion cell axon bundles that comprise the RNFL and reveals depth-dependent changes that cannot be captured by conventional OCT alone. Research AOSLO and FFOCT yielded consistent measurements of RNFB width, confirming that accurate structural information can be extracted from both modalities. On average, RNFBs were narrower at greater retinal depths and in older participants. This finding may reflect a lower axonal count with increasing age, consistent with known age-related changes in the RNFL (*24, 28*). Notably, we observed a faster age-related decrease at greater retinal depths, which may suggest a preferential early involvement of deeper layers in axonal loss. This observation also raises the possibility that the deeper portions of the RNFL are more vulnerable to mechanical forces, potentially affecting RNFBs located in these regions compared to more superficial ones. RNFB width as measured with high resolution imaging showed no significant linear relationship with the averaged RNFL thickness measured by OCT (i.e. thicker RNFL does not mean wider bundles), reinforcing the complementary value of high-resolution en face imaging in a clinical setting. Similarly, no correlation was found between RNFB width and either tortuosity or orientation at 4°N. However, a consistent boundary effect was observed for tortuosity, suggesting a geometric constraint whereby globally tortuous bundles are systematically narrower. Finally, quantification of global RNFB orientation allowed us to revisit the mathematical model proposed by Jansonius et al. (*16*), and create a Bézier curves atlas of RNFB trajectories, yielding revised trajectory equations that more accurately describe fiber paths. From a clinical perspective, these equations could be integrated into a software tool requiring only the selection of the fovea, ONH center, and ONH radius on a retinal image. The corresponding RNFB trajectories could then be displayed immediately as an overlay, enabling direct visualization of deviations from normal trajectories. In a second step, color coded orientation maps, similar to RNFL thickness maps used in clinical OCT systems, could also be generated to highlight localized angular abnormalities relative to healthy eyes. These highlighted regions could then be evaluated in further detail for bundle thinning, local orientation, and tortuosity indicators using our image processing tools.

While previous studies have primarily assessed global RNFL thickness (*7*), our approach provides depth-resolved en face structural information allowing for the quantification of bundle width.

Consistent with AO-OCT findings (*32*) showing that RNFBs are not perfectly cylindrical, bundles appeared wider at shallower retinal layers when capturing the same bundles across depths with FFOCT. When the exact same retinal location was compared, FFOCT consistently measured narrower structures than AOSLO. This discrepancy is attributable to the poorer axial resolution of AOSLO, which integrates signal contributions over an approximately 40 μm-thick layer (*33*), whereas FFOCT can distinguish shallow from deep structures thanks to its 8 μm-thick optical sections.

Regarding the relationship between age and RNFB width, a decrease with age was expected, consistent with previously reported age-related RNFL thinning (*24, 28*). Given that our cohort spanned a relatively narrow age range (19–45 years), we interpret this trend with caution. Interestingly, the decline in width was more pronounced in deeper layers, corresponding to the narrowest bundles. This suggests that those fibers may be particularly vulnerable to age-related loss, possibly due to their anatomical or mechanical properties. Such findings position FFOCT as a strong candidate for detecting early structural changes, particularly in regions where the RNFL is thinner, as it captures depth-specific information that AOSLO cannot resolve.

To connect these potential biomarkers to clinical practice, Spectralis averaged thickness maps were analysed and compared with the bundle widths measured on FFOCT and research AOSLO en face images. While these comparisons shown no significant linear relationship, we believe that linking OCT with high-resolution en face imaging will remain valuable. For example correlations between other features and information in raw OCT images could be explored, such as FPF.

Local trajectory and tortuosity quantification provided intra-bundle structural information and allowed us to investigate potential correlations with width. The boundary effect we identified indicates that tortuosity is possible only for thinner bundles.

For the characterization of global RNFB trajectories, large en face retinal montages with consistent image quality are required, and fast acquisition over a wide field of view makes Mona IV AOSLO the most suitable modality in this context. The global orientation model obtained by combining existing mathematical frameworks with the segmentation pipeline developed in this study provides a more accurate orientation map that can be further refined as additional data from healthy participants become available. This model serves two main purposes: enabling comparison between healthy and diseased eyes, and guiding the analysis of individual axon bundles without the need for manual tracing. Because it is grounded in high-resolution imaging, the segmentation accuracy supports the generation of precise, individualized orientation maps, facilitating patient-specific follow-up using devices compatible with clinical deployment. The Jansonius et al. (*16*) model was primarily built on arcuate fiber bundles as seen in color fundus photographs, and it is precisely in these regions that it performs best. The color fundus photos do not provide high contrast of the fibers close to the raphe and thus it is not surprising that the Jansonius model would not have been able to predict the true orientation of the fibers in that area since it lacked data from that location. The temporal region is known to exhibit the greatest inter-individual variability and is the least constrained by the original model, which explains why our high-resolution data lead to the most substantial refinements in this area (*34*).

In addition, the axial resolution of FFOCT could provide further information about deeper RNFB orientations. It remains unknown whether these follow the same patterns as those observed with the Mona IV AOSLO or whether deeper bundles located near the RNFL boundaries exhibit an angular shift. If such orientation shifts are present, they should be quantified and incorporated into the model and trajectory equations.

However, several limitations should be considered in this study. Both AOSLOs and FFOCT modalities require careful control of acquisition conditions to ensure repeatability between sessions. Pupil dilation and suppression of accommodation are necessary for FFOCT to achieve optimal image quality, as its beam diameter of approximately 7 mm requires a fully dilated pupil for best aberration correction. For the research AOSLO, accommodation and dilation must similarly be controlled, whereas Mona IV is less sensitive to pupil size, and a diameter of 4 mm is enough, due to its smaller beam diameter. Also, both FFOCT and AOSLO techniques are sensitive to retinal curvature (*35*) (cf Materials and Methods) which creates variability in signal homogeneity. Beyond these shared constraints, both AOSLOs are inherently limited by their optical sectioning of approximately 40 μm.

Regarding FFOCT limitations, the main source of variability in FFOCT image quality arises from signal strength. If the signal is not high enough it prevents correct tracking of the eye and adjustment of patient alignment (see Materials and Methods). Even in the case of a strong signal, there is a residual axial motion compensation error of around 10 μm rms, due to the limited rate of the stabilization control loop and motor positioning errors. It can result in multiple depths being acquired for a single nominal depth offset. Unexpected axial motion caused by changes in forehead pressure or participant movement further affects registration quality. Chinrest stability, patient positioning, and fixation target design are therefore critical. Because the eye is never perfectly stable during fixation, small lateral fixational eye movements occur continuously during acquisition (*36*). As a result, in addition to grouping images by axial position, the clustering algorithm also implicitly captures these lateral shifts caused by fixation instability. Consequently, although ideally only 11 averaged images (corresponding to the 11 applied depth offsets) would be expected, the final registered dataset contains between 11 and 55 averaged images per participant, derived from the 8000 raw images. The inter-individual variability in RNFL thickness also drives the difference in the number of registered images. Finally, although the volume is globally sorted by depth, manual verification of layer ordering from ILM to GCL remains necessary at this time.

Concerning the analysis pipeline (see Methods), bundle filtering was essential for reliable segmentation, particularly for FFOCT images which exhibited higher noise levels. The elliptical Gaussian filter enhances structures aligned along a dominant direction and is adaptable to different morphologies and image qualities. However, when blood vessels are present within the region of interest, their orientation may dominate in the Fourier domain and lead to erroneous filtering. This issue arises predominantly in AOSLO images due to the poorer optical sectioning, which makes the vessels visible when the RNFL thickness is less than approximately 40 μm, but can be mitigated by selecting smaller regions of interest or excluding images with excessive vessel coverage.

The global orientation map also retains a partial contribution from the Jansonius mathematical model, used to fill gaps where no fibers were segmented within and around the optic nerve head especially, or in vessel-affected areas, introducing a small bias in the final output.

Finally, the cohort was restricted to participants aged 19–45 years, and recruitment of older healthy volunteers will be necessary to establish broader normative values and avoid age-related bias in future comparisons with patient data, particularly as glaucoma and neurodegenerative diseases predominantly affect older populations.

Local orientation and tortuosity analyses were limited to 4°N, where bundles are relatively homogeneous. Extending this analysis to the raphe and the papillomacular bundle, which are the regions of greater architectural complexity, would require targeted protocols and larger cohorts, but could reveal whether bundle morphology is locally constrained by trajectory geometry in areas of high fiber convergence.

Each modality presented here offers distinct advantages for the early diagnosis of glaucoma, a condition known to be associated with progressive RNFL thinning (*22*). Beyond detection, a deeper understanding of the mechanisms underlying RNFB degeneration remains a critical unmet need. Fundamental questions remain open: how does degeneration propagate, why does it proceed asymmetrically despite uniform intraocular pressure distribution, and what is the relationship between vascular integrity and fiber loss? High-resolution en face imaging is uniquely positioned to begin addressing these questions at the structural level. By providing the first clinically suitable depth-resolved characterization of RNFB microstructure in healthy eyes, this study establishes the structural reference framework necessary to detect and localize the earliest signs of fiber degeneration, a question that conventional OCT, limited to global thickness measurements, cannot address. While AO-OCT remains the technique offering the highest resolution and volumetric imaging capability (*32*), FFOCT provides several practical advantages for clinical translation. This includes a smaller footprint, faster acquisition, a larger field of view, and a simplified AO correction approach due to the use of a spatially incoherent light source (*29*), making it particularly suitable when imaging time is limited or when large retinal areas must be assessed. The multimodal approach adopted here allowed us to cross-validate measurements and better define which system and which biomarkers are most suited to specific clinical contexts, while also clarifying the sources of interindividual variability in RNFB width previously reported in the literature (*37, 38*).

Future studies should investigate whether the normative biomarkers identified here are clinically relevant by recruiting cohorts including glaucoma suspects and patients at early and mid-stages of the disease. Such data would provide a framework for determining where the first structural changes appear and how fiber bundles evolve throughout the degenerative process. Do global fiber trajectories reorganise as localized losses occur? Does degeneration initiate at the superficial or deep layer of the bundles? Does reflectivity loss precede width reduction and can width reduction be measured in these modalities in the context of reflectivity loss? These questions will guide our future investigations, alongside the continued enrichment of the normative database to ensure it is representative of the broader population.

Finally, beyond retinal fiber bundles themselves, the dark spaces observed between them in en face images may correspond to Müller cell processes, which are known to occupy the interstitial space within the RNFL and form the structural basis by which the axons are packaged into the RNFBs (*7*). Whether Müller cells actively constrain bundle organisation, and whether bundle packing density reflects local Müller cell density, remains an open question that cannot be addressed with reflectance-based imaging alone. Fluorescence imaging in animal models could provide a means to directly visualise Müller cell organisation relative to fiber bundle architecture and test this hypothesis. Glial cells more broadly, including astrocytes, are known to be sensitive to mechanical forces and have been linked to collagen network disorders (*39, 40*). It has been demonstrated that AOSLO offset imaging in phase contrast, which enhances multiply scattered light, can reveal glial cells (*9*). Their potential role in modulating bundle width through mechanobiological pathways represents a complementary avenue for future investigation that is particularly relevant in the context of glaucoma, where intraocular pressure-induced mechanical stress is a central feature of the more typical form of the disease although fiber degeneration can also occur in normal-tension conditions.

This study is the first to use FFOCT to observe and quantify RNFL microstructure at multiple depths, providing a unique window into the organisation of fiber bundles that has not previously been accessible with clinical devices. By establishing normative depth-resolved biomarkers in healthy participants this study creates the conditions for investigating where and how degeneration first manifests. The algorithms developed to enhance and segment fiber bundles are applicable to any cylindrical structure in biological or non-biological images, representing a methodological contribution that extends beyond retinal imaging. Finally, the two models of RNFB trajectories we proposed can be directly used by clinicians to evaluate deviations from normal RNFB trajectories by selecting only three points: the fovea, ONH center, and ONH radius. Once glaucoma patients have been imaged and the most clinically relevant biomarkers confirmed, the analysis pipeline will be automated and integrated into a user-friendly interface designed for use by clinical personnel and non-expert users alike.

## Materials and Methods

### Imaging devices

#### Clinical SD-OCT and cSLO

The Spectralis HRA+OCT (Heidelberg Engineering, Germany), a commercial device combining confocal Scanning Laser Ophthalmoscopy (cSLO) and Spectral-Domain Optical Coherence Tomography (SDOCT), was used to obtain both en face fundus images and high-resolution crosssectional images of the retina over a field of view of 30° x 30°. The SD-OCT has a A-scan rate of 85 kHz, an optical lateral resolution of 14 μm (*41*), an axial resolution of 7 μm, and a scan depth in tissue of 1.9 mm.

### Full Field Optical Coherence Tomography retinal imager

Time-domain Full Field Optical Coherence Tomography (FFOCT) is a variant of OCT using a full-field illumination and detection scheme to acquire high-resolution en face images without scanning (*42, 43*). Owing to the use of a low coherence light source, such as an LED, interference only occurs in a narrow slice called the coherence gate, resulting in depth-resolved images of the sample. For this study, two identical custom built FFOCT retinal imaging systems were used, one at the 15-20 National Eye Hospital in Paris, and the other at the UPMC Vision Institute in Pittsburgh. They provide 5° x 5° images of the retina with 2 x 2 μm lateral resolution and 8 μm axial resolution (*29*). In this clinically-adapted implementation, FFOCT is coupled with SDOCT, allowing tracking of the retina for real-time axial motion compensation and positioning of the coherence gate at the layer of interest. Aberration correction is done using the “adaptive glasses” approach, where the wavefront is optimized in a sensorless manner thanks to tunable transmissive elements in front of the eye, using SDOCT signal as a surrogate for FFOCT image optimization (details in Mece et al. (*29*)). In this study, two lenses were combined. Focusing on the RNFL is achieved with an electrically tunable lens (ETL, Optotune, Switzerland) with 16 D defocus correction range. Higher order aberrations are corrected by a multi-actuator adaptive lens (MAL, Dynamic Optics srl, Italy). Special attention was given to the chinrest and fixation target in this study, as the technique is particularly sensitive to axial motion. To address this, we designed a custom-made, rigid chinrest equipped with adjustable pods on each side to maintain the forehead stable. In addition, we created a fixation target outside the system to extend the imaging FOV we had access to, based on a mirror located in front of the fellow eye onto which a point is projected.

### Adaptive Optics Scanning Laser Ophthalmoscopy

Adaptive Optics Scanning Laser Ophthalmoscopy (AOSLO) is a high-resolution imaging technique providing en face images. In this study we use two AOSLO devices. One is a research AOSLO from the University of Pittsburgh (*44, 45*) and the other a commercial AOSLO (Mona IV, Robotrak Technologies Inc., Nanjing, China) (*46, 47*). Data were acquired using these two AOSLO system configurations depending on device availability during the study period.

The AOSLO at the University of Pittsburgh is a custom built setup that gives 1.5° x 1.5° in-vivo cellular-level images with a lateral resolution of 2-3 µm and an axial resolution of approximately 40 µm (*33*) by the use of a closed-loop AO system to correct the wavefront aberrations of the human eye. It uses a supercontinuum light source for illumination (720 nm light with a 20 nm bandwidth was used for these experiments) and a 909 nm laser diode for wavefront sensing.

The commercial AOSLO has a lateral resolution around 2 µm and an axial resolution around 40 µm. The system allows 1.2°, 2.4° and 5° FOV and uses a super-luminescent diode for imaging at 840 nm. Increasing the field of view while keeping a constant pixel number results in a larger pixel size, thus increasing the µm per pixel value from approximately 0.7μm/pixel (1.2° FOV) up to 3μm/pixel (5° FOV), for a 1024 x 1024 sampling.

### Participants

14 healthy volunteers from 19 to 45 years old, of which 8 were female participated in these studies. Table 2 lists their age and the imaging modalities used. For all of them only the right eye was imaged.

**Table 2.**
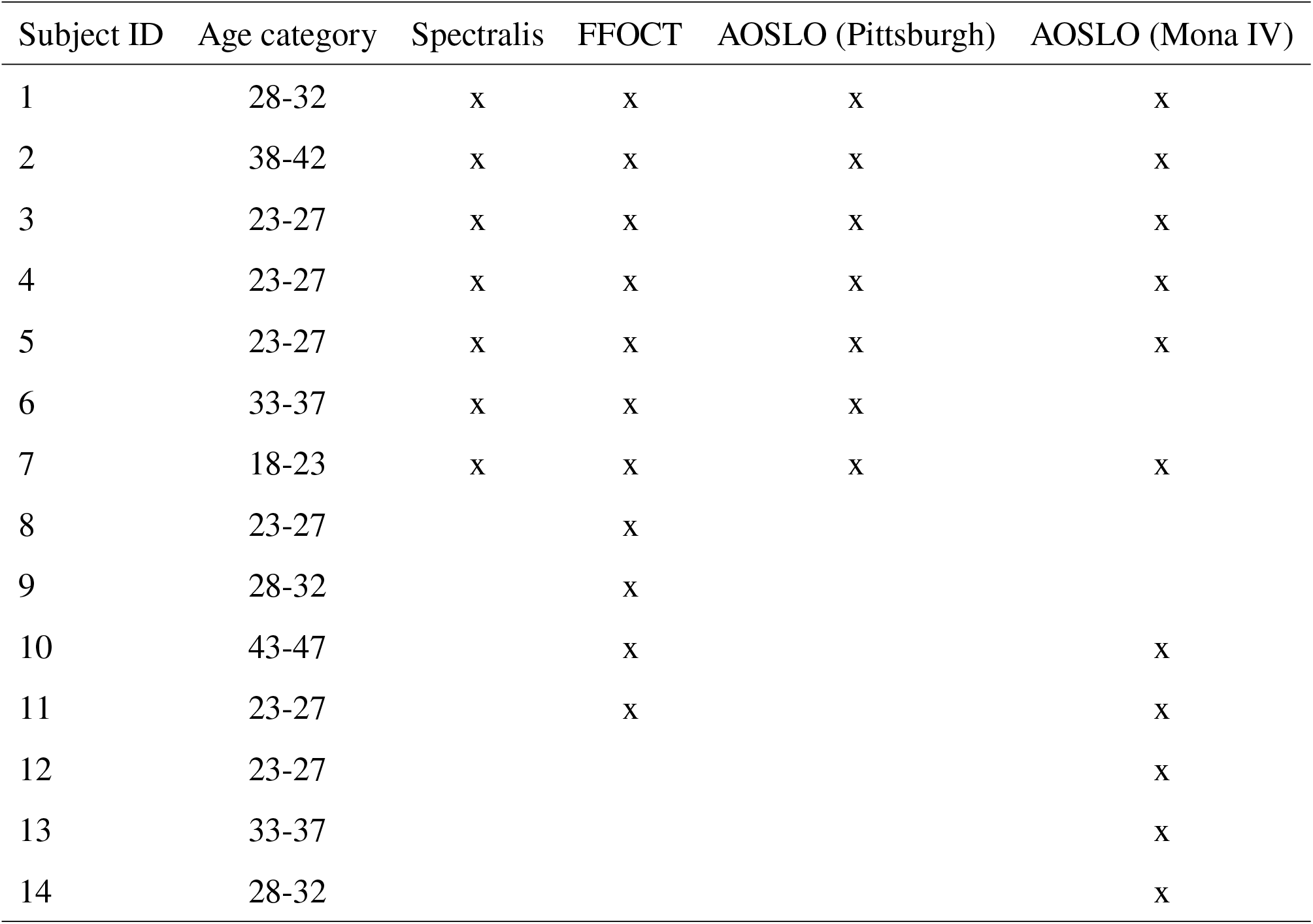
Subjects information and imaging modalities used. Each row corresponds to a subject. The table reports age and ssthe imaging modalities acquired: Spectralis OCT, FFOCT, and AOSLO systems (Pittsburgh and Mona IV). An “xs indicates that the modality was acquired.

The study was conducted according to the Declaration of Helsinki and approved by the Ethics Committee in Paris (IDRCB number: 2019-A00942-55, CPP Sud Est III: 2019-021-B) and the Institutional Review Board of the University of Pittsburgh. Written informed consent was obtained from all participants following a detailed explanation of the associated risks, provided both verbally and in writing. Before image acquisition, light levels for imaging were measured, for FFOCT and AOSLO, to verify that they remained below the maximum permissible exposure specified by the ANSI and ISO standard guidelines.

### Image acquisitions

Pupillary dilation was achieved by administering one drop each of phenylephrine hydrochloride (2.5%) and tropicamide (1%) approximately 15 minutes prior to imaging. OCT images were acquired at two different locations using the Spectralis: one measurement was centered on the fovea (0°, 0°) and two were centered on the optic nerve head. Both research AOSLO and FFOCT images were acquired at 4°N in the RNFL. 5° FOV Mona IV image tiles were acquired and montaged to create a wide FOV of 14° by 26°, from 12° N to 10° T and from 6° superior to 3° inferior.

### Spectralis Protocol

Three measurements were performed on 7 participants (participant #1 to #7) in the glaucoma mode setting of the Spectralis. First, two dense OCT vertical scans with 30 μm spacing, one centered on the fovea, the other on the ONH. Then, a thickness profile of the RNFL around the ONH. Finally, two infrared cSLO fundus photos, one centered on the macula, the other on the ONH.

### FFOCT Protocol

FFOCT retinal imaging was performed on 11 healthy participants. 4 were imaged in Paris, 6 in Pittsburgh and 1 (#4) at both sites using identical custom-built systems. First, we positioned the FFOCT coherence gate at the Inner Limiting Membrane (ILM) utilizing the SDOCT B-scan and A-scan. The coherence gate functions as a depth filter, since recorded interferences can only occur in structures within a thin slice at the corresponding depth. Here, the slice is 8-μm thick. The SDOCT signal is then used to control the position of the FFOCT coherence gate, enabling acquisition at *N* = 11 depths with a spacing Δ*z*, ranging from ILM +10 μm to ILM −40 μm. The corresponding FFOCT slices are illustrated in blue on the SDOCT B-scan (Figure 5), forming the reconstructed FFOCT volume. In our protocol Δ*z* = 5 μm to ensure signal continuity. This resulted in a stack of 8000 FFOCT en face images (around 727 per depth position). We refer to this as the multi-depth approach. A high number of images is required due to our sensitivity to axial eye motion that leads to frame loss, and to the low SNR of individual FFOCT images, which must be averaged through our registration process to obtain high-contrast images. Note that the curved shape of the RNFL in depth can prevent the flat coherence gate from covering the entire field at a given depth offset (*35*) resulting in locally missing regions. These are however recovered by imaging at adjacent depth offsets.

**Figure 5.**
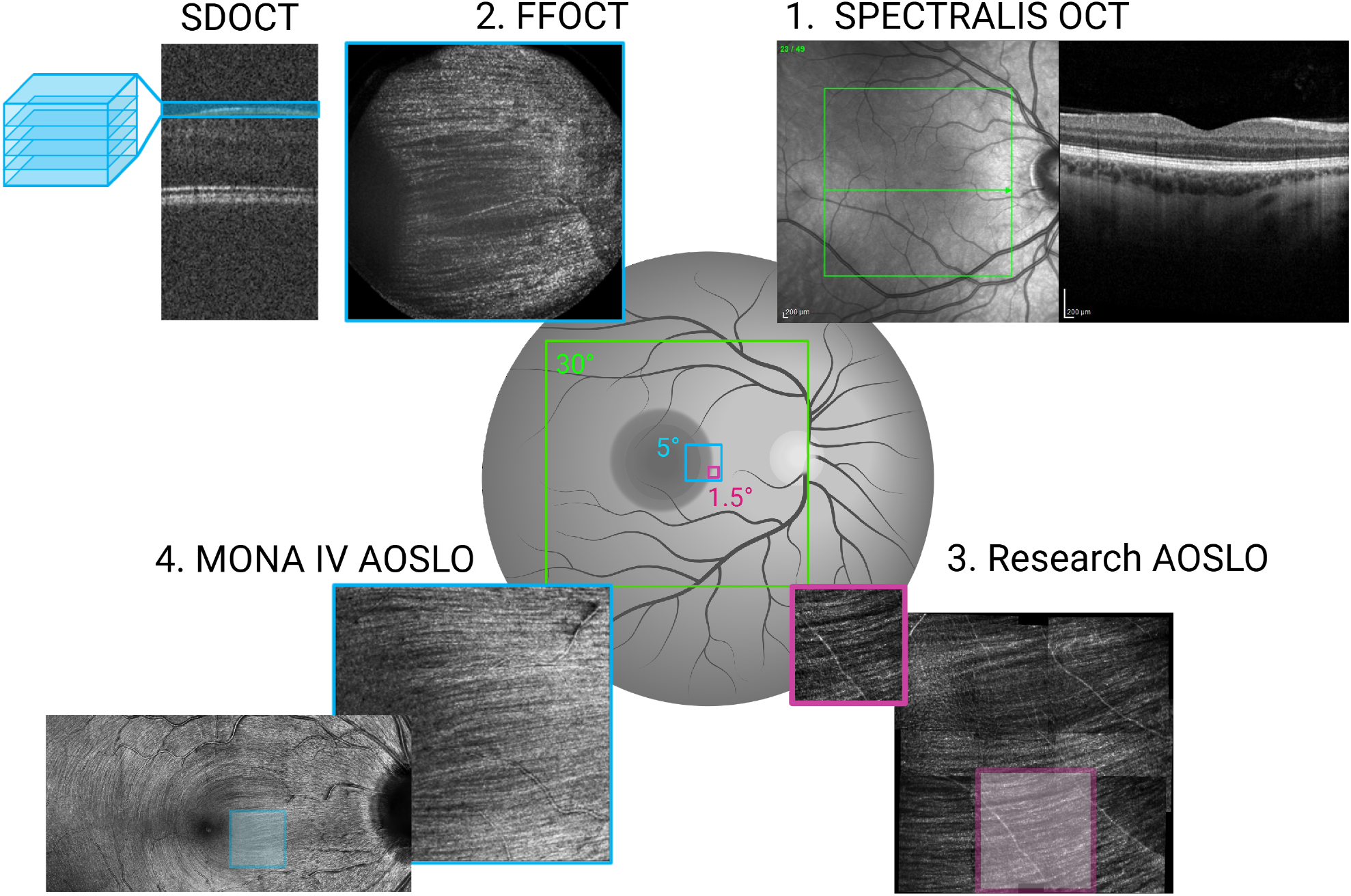
Multimodal image acquisition protocol. The figure summarizes the acquisition workflow across OCT and AOSLO modalities. (**A**) Spectralis OCT acquisition consisting of three repeated measurements. (**B**) FFOCT multi-depth imaging protocol guided by SDOCT. The SDOCT signal is used to control the position of the FFOCT coherence gate, enabling acquisition at *N* = 11 depths with a spacing Δ*z*, ranging from ILM +10 μm to ILM −40 μm. The corresponding FFOCT slices are illustrated in blue on the SDOCT B-scan, forming the reconstructed FFOCT volume. (**C**) Research AOSLO acquisition of nine images centered around 4° nasal. (**D**) Commercial AOSLO acquisition composed of 44 image tiles spanning from 12° nasal to 10° temporal, and from 6° superior to 3° inferior.

### AOSLO Protocol

With the research AOSLO from Pittsburgh, retinal imaging was performed on 7 healthy participants in Pittsburgh immediately following FFOCT imaging. 9 videos of 30 seconds were acquired with 1° step between acquisitions to create a montage of 3° x 3° centered at 4°N. At all times we aimed to maintain focus on the RNFL.

With Mona IV, the commercial AOSLO, we generated a wide field fundus mosaic. We imaged 10 participants from 12° Nasal to 10° Temporal in 2° steps and from 6° Superior to 3° Inferior by 3° steps. We used a 5° FOV and adjusted the focus at each position to maintain high contrast of the axon bundles.

### Image Processing

The image processing workflow, implemented in Python 3.11.9, MATLAB (R2023b; The MathWorks Inc., Natick, MA) and Fiji ImageJ (*48*) (v1.54p, NIH, USA) is presented in Figure 6. The Spectralis commercial software interface (HEYEX v2.6.7, Spectralis Software v7.2.4-US, Heidelberg Engineering, Germany) provided thickness maps, segmentation of the RNFL and evaluation of the ONH RNFL thickness according to the European Descent database (2019).

**Figure 6.**
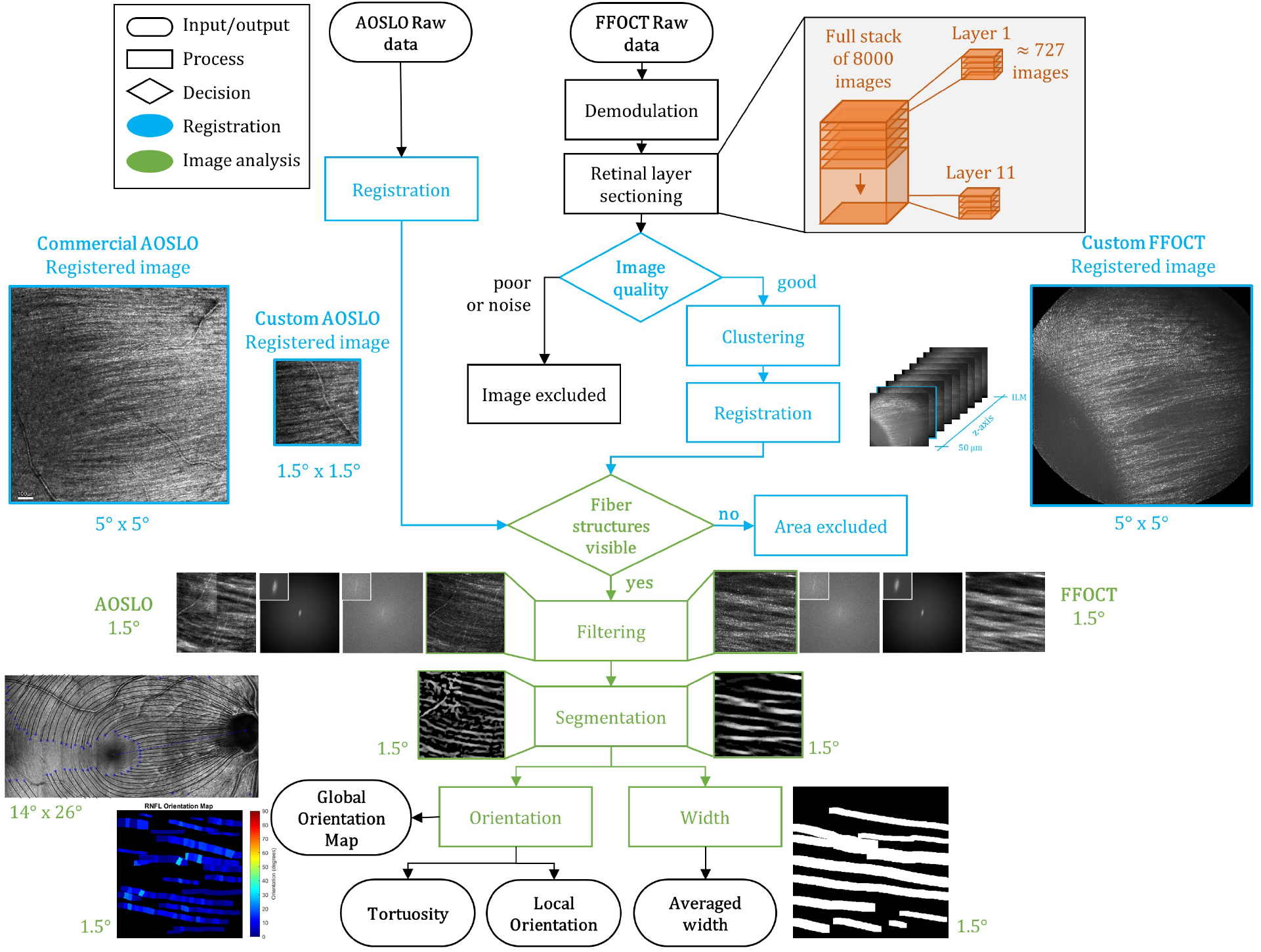
Processing workflow for multimodal RNFL analysis. The pipeline summarizes pre-processing, registration, and quantification steps applied to FFOCT and AOSLO data. (**A**) FFOCT preprocessing: raw data undergo two-phase demodulation to generate tomographic images, which are divided into 11 sub-stacks corresponding to discrete depths *z* = *n*Δ*z*, with *n* ∈ {0, …, 10} and Δ*z* = 5 μm. Noisy frames are excluded. (**B**) Reference selection and lateral registration: each sub-stack is averaged in groups of 20 frames to reduce noise. Candidate reference frames are selected from local intensity maxima and clustered based on correlation, yielding 1–5 reference images per depth. Lateral motion is corrected using a phase-correlation approach (*49*), followed by frame averaging. (**C**) Depth alignment: images across depths are registered using template matching in ImageJ, producing a multi-depth FFOCT stack. (**D**) AOSLO processing: image registration is performed using REMMIDE (*51*), followed by RNFL enhancement with an elliptical filter. (**E**) Bundle segmentation and quantification: RNFL bundles are segmented to extract a skeleton, which is then used to estimate bundle width and orientation.

For FFOCT, pre-processing, exclusion of noisy images and selection of references by clustering are specific to the FFOCT multi-depth approach as pictured in Figure 6. We adapted the image processing steps detailed in Mecê et al. (*49*) to our RNFL imaging protocol. The previous algorithm was developed for photoreceptors and included: 1) image normalization by dividing each frame by its mean intensity; 2) 2-phase demodulation by subtracting consecutive images and computing their absolute value; 3) frame rejection for frames acquired during blinks, micro-saccades, or those with low SNR due to poor phase modulation; 4) image registration to correct both translation and rotation using a phase correlation method described in Mecê et al. (*12,50*); and 5) image averaging. Here for the multi-depth approach, steps 1, 2, 4 and 5 are preserved, and new steps are introduced. Instead of step 3, a new image selection is performed, and before step 4 we introduce a new clustering approach to find the registration references, the details of which is describe in the Supplementary Material. Registration on the research AOSLO was done with REMMIDE, see Zhang et al. (*51*) for details. Finally, Mona IV AOSLO images were registered and montages automatically generated with the manufacturers built-in software (MonaPro v1.00.04.251030). After registration, the processing steps for the biomarker extraction and analysis are applied to both AOSLO and FFOCT registered images following the same workflow.

### Coarse Registration

The coarse registration of FFOCT images follows the Mece et al. (*50*) approach where a custom-built normalized cross correlation algorithm registers the useful images at a sub-pixel scale. It associates each frame to its correct reference as well as correcting for lateral motions. The resulting stacks are then averaged to give one frame per depth. These are then aligned with the `template matching` function in ImageJ providing a multi-depth stack from 11 to 55 images from the ILM to 50 μm deeper in the RNFL.

AOSLO confocal image sequences acquired with the previously described research AOSLO imaging system (*44, 45, 52*) were desinusoided using a published method (*53*) to correct the sinusoidal distortion introduced by resonant scanning, then registered using a strip-based algorithm (*51*) to correct for eye motion, and subsequently averaged to generate the final images.

As stated previously, MonaPro software (v1.00.04.251030) provided the registered images and montage directly from the Mona IV AOSLO.

### Fiber bundle enhancement

In order to analyse structures of the RNFL and better visualize the fiber bundles we developed a frequency-oriented elliptical Gaussian filter based on a Fourier space approach. This filter enhances the bundle structures by convolving each sub-image with its respective elliptical oriented filter. To achieve reliable results, we first need to identify a region of interest (ROI) where the fiber bundles are visible. For this study we decided to crop FFOCT and AOSLO images to 500 x 500 pixels to have comparable results because the FOV of those modalities differs from 5° ≈ 1.5*mm* for FFOCT to 1.5° ≈ 0.45*mm* for AOSLO. Once the ROI is selected, our MATLAB algorithm will divide it into 4 blocks and apply the filtering to every individual block. The algorithm can be divided into 5 main steps : 1) Compute 2 dimensional Fourier Transform (FFT2) of the image 2) Extract the brightest points (from 3 to 200 points), 3) Fit an ellipse around the points 4) Create an elliptic Gaussian 5) Apply the elliptical Gaussian filter to the FFT2 image before coming back to direct space.

After dividing the input ROI into four, the first step is to move into the Fourier space by applying a Fast Fourier Transform. Then we begin the second step which is the heart of the filter: finding the brightest points to fit to an ellipse. To accomplish this, we apply an adaptive threshold defined as threshold = (2 + *i* × 0.1) × mean(|*I*|). As long as the brightest points are outside the range 3 < *n*_brightest points_ < *max points* where *max points* = 11, this threshold increases progressively with *i*. However, after 1000 iterations if no solution is found we consider we cannot perform the elliptical filtering and will then apply a classic Gaussian filter. Once the points are extracted, we perform an outlier removal step, by setting an upper limit using the 3*σ* rule where any point further than *mean* + 3*σ* is considered too far away. Then we compute the coefficients of the general conic equation with the bright points. If an ellipse is found we move to step 4; otherwise we retry with *max points* + 1. If we reach *max points* = 200 and no ellipse is found once again we consider we unable to find a suitable filter and will apply a classic Gaussian filter with 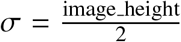. Now at step 4, we fit an ellipse to the points. Then we create an elliptical Gaussian filter defined as:

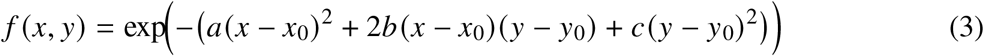

with

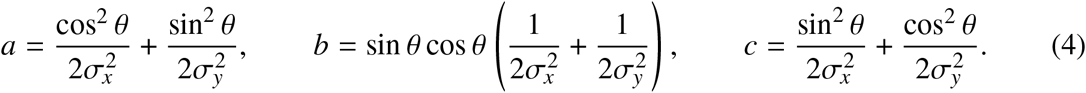

The ellipse is centered at (*x*_0_, *y*_0_). Its amplitude decreases smoothly from the center following an elliptical contour. The parameters *σ*_*x*_ and *σ*_*y*_ control the width of the ellipse along the horizontal and vertical axes, respectively, while *θ* defines the orientation of the ellipse in the image plane. Finally, a scaling factor (*fac*) adjusts its overall size. For this filter *σ*_*x*_ = 3, *f ac* = 3, and *σ*_*y*_ and *θ* are defined by our ellipse fitting. The final filter is a combination of the elliptical filter and a basic Gaussian with 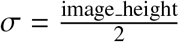 such as Final Filter = 0.35 × GaussianFilter + 0.65 × EllipticalFilter. We can then convolve the final filter to the FFT2 and go back to the direct space. This filtering enhances the bundle structures as shown in Figure 8.A. and enables measurement of the bundles.

### Bundle Segmentation

Segmentation is performed on the filtered images and is processed per block. The fiber bundles were isolated from the background by applying Frangi vessel segmentation (*54*) followed by morphological transformations to create the skeletons of their paths.

For the Frangi segmentation we set *σ* = 8, the standard deviation in each direction of the Gaussian kernel, which determines the width or scale of the Gaussian. A higher sigma value will highlight bundles of a wider diameter, and vice versa. *β* = 0.7 controls the sensitivity of the line filter to the blobness measure and is set to 0.5 in the paper by Frangi et al. A higher beta value will give more weighting in the output image to pixels containing blob-like features, and vice versa. Then we binarize the result and apply morphological operations as follows. First, we eroded the image with a line of 20 pixels oriented according to *θ_ellipse* we found with our filter to separate the bundles from the background that contain vessels. Then we dilate the image with a rectangle of 4 by 20 pixels, oriented by *θ_ellipse*, to reconnect the bundles together. We then skeletonize which gives a 1-pixel-wide skeleton, while removing branches shorter than 30 pixels. We clean up the skeleton by removing all connected components that have fewer than 40 pixels. The last step is to reconnect the block results. We connect the blocks by dilating them again with a line at successive angles corresponding to the average orientation between neighbouring blocks. We then skeletonize and clean the result again by subtracting long-branch skeletons greater than 1200 pixels from shorter ones that are less than 60 pixels.

If our filtering fails and does not provide *θ_ellipse* or if we want to use this segmentation on unfiltered images, we use the exact same pipeline but with *θ*_*radon*. It computes the angle by applying the Radon transform method to the binary image. The Radon transform determines the dominant orientation in an image by projecting it over a range of angles and selecting the angle where the summed projection intensity is maximal. More generally, the choice between *θ*_ellipse_ and *θ*_radon_ is made adaptively based on local image quality and structural content, as their performance varies across regions. The segmentation results in a skeleton corresponding to the RNFL bundle (RNFB) paths.

### Width extraction

This step is implemented to automatically measure the width of the RNFB through the different depths of the RNFL, ensuring consistent and precise data extraction. The skeleton of the axon bundle paths from the segmentation step is the input to a custom MATLAB script for the width measurements. The paths defined by the skeleton were used to guide the measurement of the width of each axon bundle from FFOCT images. The procedure we employed was to use a sliding window to move along the path of each bundle, one pixel at a time, and at each point to extract the intensity profile of a line orthogonal to the bundle path from the FFOCT image. Cross-sectional intensity profiles for each bundle segment were then averaged. The averaged cross-sectional profiles had distinct minima on each end; the distance between them was taken to represent the average width of that bundle segment. Each bundle path was evaluated sequentially with a sliding analysis window. A 10 μm wide analysis window was used for all measurements (any bundle path whose long axis was shorter than the analysis window was excluded from measurement). MATLAB functions `bwlabel` and `regionprops` with the arguments ‘Orientation’ and ‘Centroid’ were used to determine the center of the bundle path segment and its angle. These parameters permitted the determination of a line that ran orthogonal to the bundle. The intensity profile of this line was then extracted from the FFOCT image using the MATLAB function `improfile`. To ensure that it was longer than the width of the bundle, the length of each line segment extracted from the FFOCT image was set to be 6 pixels longer than the coarse width estimate made during the bundle path calculation. All intensity profiles for each individual bundle path were averaged and the distance between the first and last minima of the averaged profile was taken to represent the average width of that bundle. This process was repeated for all bundles in each FFOCT image. If no minima were found, or the average width result is greater than 85 pixels or less than 12 pixels, we consider that there is no valid measurement possible for that bundle and set the width to NaN. To visualize the result of the width measurements, we then dilate the skeleton to its measured averaged width (Figure 8.B.).

### Layer Thickness

We extracted the RNFL thickness from the Spectralis by generating the RNFL thickness heat-map. Only the average thickness per zone (see the diagram in Figure 7) can be extracted using the built-in software of the Spectralis.

**Figure 7.**
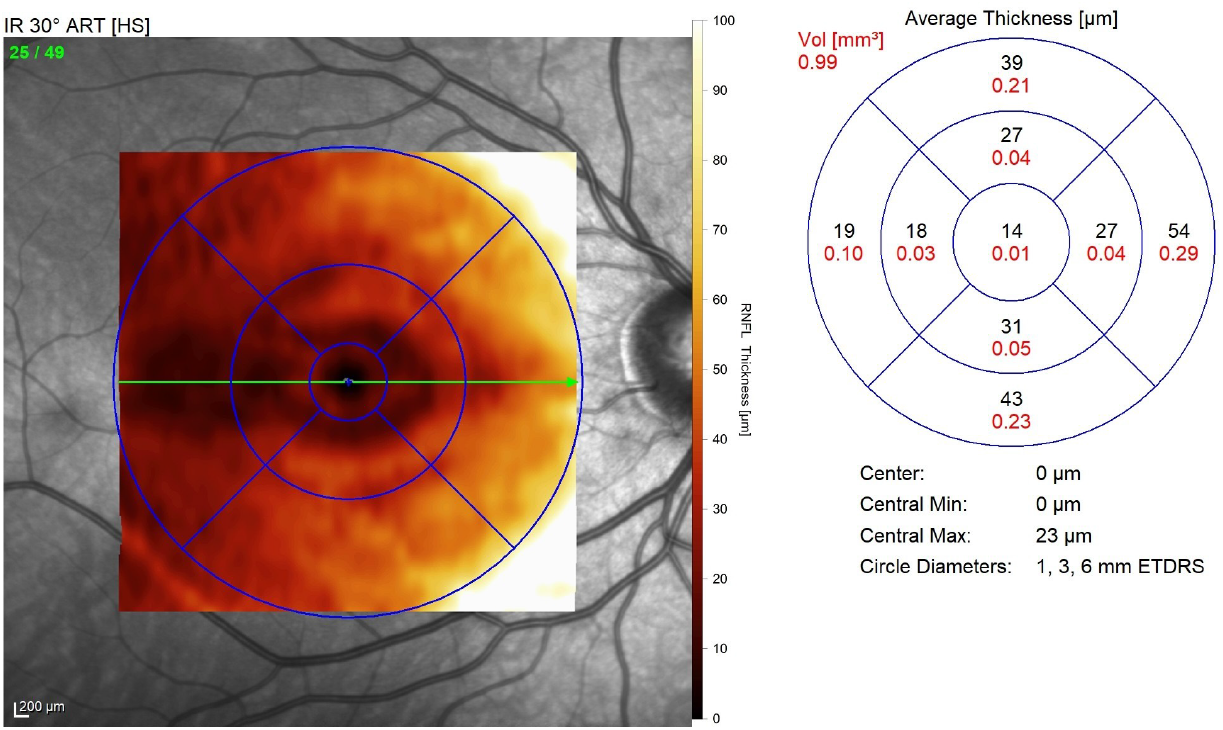
RNFL thickness heat map derived from Spectralis OCT. The heat map represents RNFL thickness measurements extracted from Spectralis OCT images. Averaged thickness values for each sector of the diagram are displayed on the right in μm. Data are shown for participant #6. The red values are the volumetric thickness in *mm*^3^ and the black values the averaged thickness in μm

### Local orientation

RNFL local orientation was quantified from skeletonized imaging data (AOSLOs, FFOCT) using a custom-developed sliding window algorithm implemented in MATLAB. Individual RNFL fiber bundles were first identified as connected components using standard binary labeling (`bwlabel` function). Fiber endpoints were identified with `bwmorph(‘endpoints’)`, and pixel coordinates were ordered by tracing the path from one endpoint through successive 8-connected neighboring pixels. Each detected fiber was subsequently divided into non-overlapping segments using a sliding window approach with a window size of 20 pixels.

For each segment, local fiber orientation (*θ*) was calculated using principal component analysis (PCA) of segment pixel coordinates (Figure 8.C.). PCA identifies the main direction of the fiber segment, and the orientation angle is computed from this direction. Raw segment orientation was then computed as 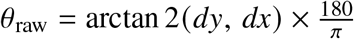 for right eyes and 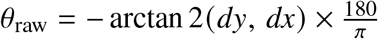 for left eyes, where (*dx, dy*) is the vector. The orientation calculation preserved directional infor-mation, yielding signed values ranging from −90° to +90°, where negative values indicate fibers ascending from left to right and positive values indicate descending trajectories. This method enables the differentiation of superior versus inferior arcuate bundle patterns, which exhibit opposite curvature directions around the optic disc. Bundles will be expected to have a different orientation depending on the eye we look at, due to the mirror symmetry between the eyes, with fiber bundles supero-temporal to the ONH ascending in right eyes and descending in left eyes.

**Figure 8.**
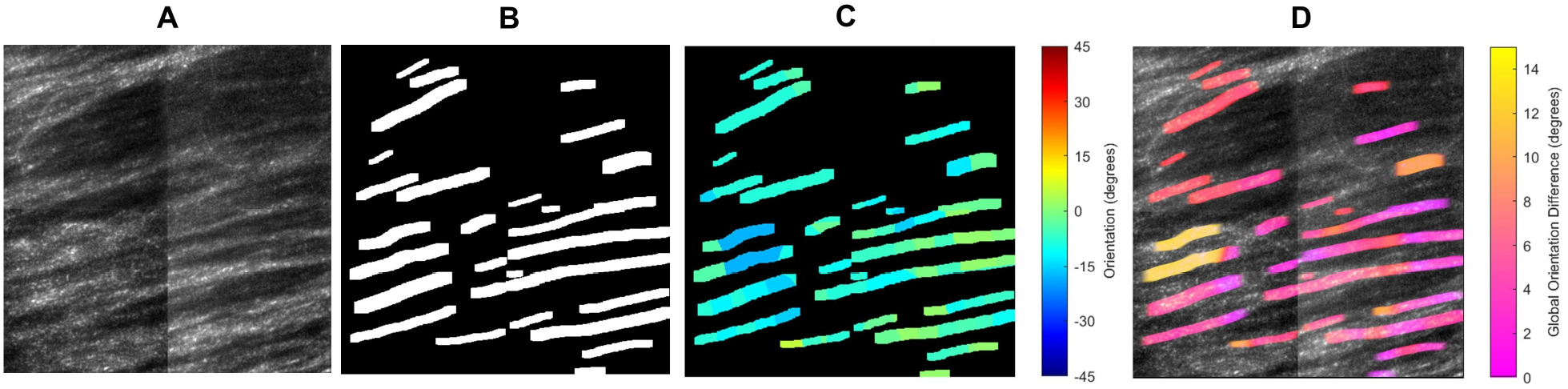
Quantitative analysis pipeline for RNFB characterization. The figure illustrates the extraction of structural features from FFOCT images. (**A**) Filtered FFOCT image. (**B**) Map of averaged RNFB width. (**C**) Map of local RNFB orientation. (**D**) Map of local trajectory differences intra-bundle to quantify local fiber smoothness.

Due to random amounts of rotation being present in the cSLO images, raw orientation values were corrected for the anatomical offset between the optic disc and fovea using patient-specific disc-fovea angles (DFA). DFA values were calculated from Spectralis OCT SLO fundus images using a two-step approach. First, the optic disc and foveal centers were automatically detected using the open-source SLOctolyzer software (*55*). This software used deep learning-based segmentation for disc localization and brightness-based detection for foveal identification. Second, the DFA was computed from the extracted pixel coordinates (*x*_disc_, *y*_disc_) and (*x*_fovea_, *y*_fovea_) using the four-quadrant arctangent function (atan2): 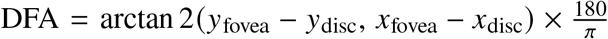, which yields the angle in degrees accounting for all quadrants. The anatomical correction was applied uniformly as *θ*_corrected_ = *θ*_raw_ − DFA for both eyes, ensuring that fibers running parallel to the fovea-disc axis were represented as approximately 0° (horizontal). This correction enables anatomically meaningful comparisons across participants and eyes by standardizing measurements to a common anatomical reference frame.

To assess local fiber smoothness and detect regions of abnormal curvature, segment-to-segment orientation differences were calculated as Δ*θ* = |*θ*_*i*+1_ − *θ*_*i*_ | for each pair of consecutive segments within the same fiber, where *θ*_*i*_ and *θ*_*i*+1_ represent the corrected orientations of adjacent segments (Figure 8.D.).

Figure 8 shows the results of the different quantifications. After selection and filtering of an ROI (Figure 8.A.) we visualize the average width measure per bundle by dilatation (Figure 8.B.) and the local orientation by a colorbar from −45° to 45° (Figure 8.C.). Finally, we highlight local orientation deviations per bundle (Figure 8.D.) from no changes in pink to 15° deviation in yellow.

### Global orientation

RNFL global orientation was quantified from Mona IV skeleton montages on 9 participants. We estimate global trajectories of the RNFB over a large FOV of 14° by 26°. There are two goals here: first, to automatically reconstruct RNFB traces from high-resolution images, and second, to propose a refined model of RNFB trajectories.

We followed Schwarzhans et al. (*17*) method but instead of using Polarization-sensitive optical coherence tomography we used AOSLO images provided by Mona IV. We started by manually selecting the fovea center, ONH center, ONH radius and raphe axis which is defined as the horizontal meridian through the fovea that separates the superior and inferior RNFL. Then, as a prior for our trace model, we used an interpolated orientation map (*OM*_*M*_) derived from the mathematical model established by Jansonius et al. (*16*), which fits the dimension of the input image and takes into account fovea and ONH positions. The other contribution to the model is the skeletonized image data from our previously described algorithm. We first cleaned the RNFB skeleton by excluding vessel segments to then estimate the local orientation (angle) by looking at nearby 15 pixels and performing a local principal component analysis. From the calculated angles, we remove those that deviated by more than 20° from the local mean orientation computed within a 100-pixel window. We finally mapped the filtered angles onto an image grid and interpolated them to obtain an orientation map (*OM*_*S*_). We left undefined regions as NaN. We then combined orientation data into a final orientation map OM. The Jansonius model contribution (*W*_*M*_) was dominant near the disc (within one disc radius) and decayed quadratically to zero at six disc radius, while the skeleton contribution (*W*_*S*_) was complementary as the final OM is equal to:

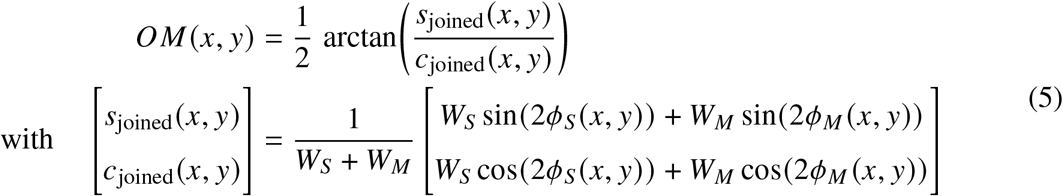

After obtaining OM, we smooth it with a Gaussian filter. As in Schwarzhans et al. (*17*) any RNFB trajectory can be traced by selecting an initial point (*x, y*) and iteratively moving a fixed step size *r* in the local fiber direction. The next position (*x*^’^, *y*^’^) is obtained by advancing *r* along the orientation given by the orientation map *OM*(*x, y*), as:

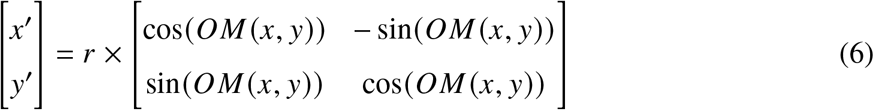

We quantified the angular difference (in degrees) between the mathematical model of Jansonius (*16*) *OM*_*M*_ and our interpolated segmentation map *OM*_*S*_, reported globally and spatially across a 100×100-pixel grid expressed in retinal coordinates (see Figure 4.C.). We also divided the image into sectors: temporal, inferotemporal, superonasal, inferonasal and superotemporal. We computed the mean error for each of them to quantify where the model differed from the segmentation map the most. The resulting sector errors were compared using a Kruskal-Wallis test, with Bonferroni-corrected post-hoc comparisons when significant (*α* = 0.05) (cf Table. 1). We then computed an inter-subject 2D map of the mean angular error expressed in retinal coordinates. (cf Figure 4.D) to understand where the model differs the most from our segmented images in our cohort.

Finally, to reach our second goal, from the individual orientation maps generated for each participant, we created our own equations of RNFB trajectories for a FOV of 14° by 26°. To do so, for each participant, we choose to generate retinal RNFB traces based on 3rd degree Bézier curves which are driven by four control points (*P*_0_, *P*_1_, *P*_2_, *P*_4_) such as:

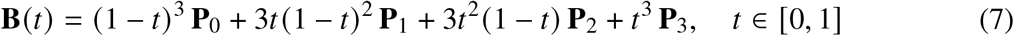

We defined 3400 starting points (**P**_0_) located around the macula, along the image borders, and near the raphe. The end points (**P**_3_) were defined on the radius of the ONH. We then estimated the intermediate control points (**P**_1_, **P**_2_) such that the Bézier curve **B**(*t*) best matched the orientation data (**q**_*i*_). The parameter *t*_*i*_ represents the relative position along a bundle path and was defined as the normalized arc-length position of point **q**_*i*_. Here is a summary of the control point definitions:

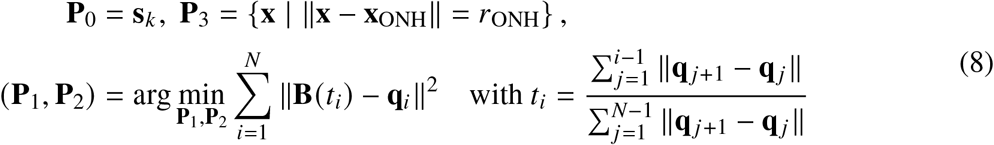

We then used two approaches to combine RNFB trajectories and find new equations. The first one was based on Bézier curves and the second on Jansonius trajectories. The goal of the Bézier curves approach was to find the best fit for our segmentation and propose final equations that are as close as possible to the traces we measure on our cohort. The goal of expressing the curves as Jansonius trajectories was to refine the existing model to better fit the observed RNFB trajectories.

For the Bézier curves approach, after expressing each participant’s curves in a common coordinate system, we computed the median angular trajectory for the fiber bundle originating from a given starting point. We then applied spline smoothing to enforce spatial regularity and balancing fidelity to the median trajectory with curvature minimization. This resulted in a set of parametric Bézier curves defined at starting points distributed across a 14° × 26° FOV. From these curves, a dense orientation map was generated by extracting local tangent directions along the trajectories and interpolating them over the image domain. This approach enabled the generation of RNFB trajectories in new images without requiring image segmentation (cf. Figure 4.F.).

For the fine-tuning of Jansonius trajectories, defined as 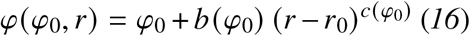, the parameters *b*(*φ*_0_) and *c*(*φ*_0_) were adjusted within a normalized spatial framework. The original Jansonius model describes retinal trajectories in visual degrees and assumes a fixed scaling between the optic nerve head and the fovea. In the present formulation, distances were normalized using the optic disc–fovea distance (DOF) measured directly on each image. The radial coordinate *r* was therefore replaced by the normalized coordinate 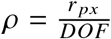, where *r* _*px*_ is the radial distance in pixels and *DOF* is the distance between the optic nerve head center and the fovea. This normalization accounts for anatomical variations between eyes while preserving the mathematical formulation of the trajectories. We first expressed each participant’s curves in a common coordinate system. We then applied a weighted non-linear least-squares fit (`lsqcurvefit`, MATLAB), with *c* set as a free parameter. Only the most distal points (*r* ≥ 98th percentile of *r*) were retained, and a *r*^80^ weighting was applied to prioritize locations far from the ONH, where the Jansonius model showed the greatest deviation from RNFB trajectories. Seeds with a weighted *R*^2^ < 0.5 were excluded from the analysis. We then modeled *c*(*φ*_0_) with a hyperbolic tangent function in the superior region and with a combination of two hyperbolic tangent functions in the inferior region. A second fit was performed to estimate *b*(*φ*_0_): the median value of *b* was computed within sliding 5° bins, followed by an exponential hyperbolic tangent fit. Seeds with a weighted *R*^2^ < 0.9 or an angular error at the ONH > 20° were excluded, yielding four final parametric functions for *c* and *b* (one per parameter per hemisphere). A valid fit for *c*(*φ*_0_) was obtained over [105°, 180°]∪ [−180°, −105°], and for *b*(*φ*_0_) over approximately [140°, 180°]∪[−180°, −105°]. The original Jansonius model was retained for the intermediate range [−104°, 104°] to enable the generation of a dense orientation map. Similarly to the Bézier approach, we were able to generate RNFB trajectories in new images without requiring image segmentation (cf. Figure 4.H.).

## Funding

This research was supported by the European Research Council OPTORETINA [#101001841], Région Ile de France Sésame 4Deye, IHU FORESIGHT project [ANR-18-IAHU-0001], ANR BRAINS [ANR-22-CE19-0010-01], and ERC [101221080].

## Author contributions

C.C. and M.B. conceived the experimental setup, based on an original idea from P.M. and K.Gr. C.C., M.B and K.Gu. collected the data. C.C. developed the processing algorithms and analysis pipeline, with contributions from E.A.R. for the RNFB width algorithm and K.Gu. for the local orientation algorithm. C.C. analysed the results. K.Gu., K.Gr., M.B. P.M. and E.A.R. provided revisions for the manuscript and all authors reviewed the manuscript before submission.

## Competing interests

M.B., P.M. and K.Gr have patents concerning FFOCT. The authors declare no other competing interests.

## Data and materials availability

The codes can be provided by C.C. pending scientific review and a completed material transfer agreement. Requests for the codes should be submitted to.

## Notes

### Funding Statement

This research was supported by the European Research Council OPTORETINA [#101001841], Region Ile de France Sesame 4Deye, IHU FORESIGHT project [ANR-18-IAHU-0001], ANR BRAINS [ANR-22-CE19-0010-01], and ERC [101221080].

### Author Declarations

The study was conducted according to the Declaration of Helsinki and approved by the Ethics Committee in Paris (IDRCB number: 2019-A00942-55, CPP Sud Est III: 2019-021-B) and the Institutional Review Board of the University of Pittsburgh. Written informed consent was obtained from all participants following a detailed explanation of the associated risks, provided both verbally and in writing.

## References and Notes

1. A. Petzold, et al., Optical coherence tomography in multiple sclerosis: a systematic review and meta-analysis. The Lancet Neurology 9 (9), 921–932 (2010), doi:10.1016/S1474-4422(10)70168-X, https://www.sciencedirect.com/science/article/pii/S147444221070168X.

2. H. V. Danesh-Meyer, H. Birch, J. Y.-F. Ku, S. Carroll, G. Gamble, Reduction of optic nerve fibers in patients with Alzheimer disease identified by laser imaging. Neurology 67 (10), 1852–1854 (2006), company: Lippincott Williams & Wilkins Distributor: Lippincott Williams & Wilkins Institution: Lippincott Williams & Wilkins Label: Lippincott Williams & Wilkins, doi:10.1212/01.wnl.0000244490.07925.8b, https://www.neurology.org/doi/10.1212/01.wnl.0000244490.07925.8b.

3. Y. Lu, et al., Retinal nerve fiber layer structure abnormalities in early Alzheimer’s disease: Evidence in optical coherence tomography. Neuroscience Letters 480 (1), 69–72 (2010), doi:10.1016/j.neulet.2010.06.006, https://linkinghub.elsevier.com/retrieve/pii/S0304394010007391.

4. J.-g. Yu, et al., Retinal Nerve Fiber Layer Thickness Changes in Parkinson Disease: A Meta-Analysis. PLoS ONE 9 (1), e85718 (2014), doi:10.1371/journal.pone.0085718, https://dx.plos.org/10.1371/journal.pone.0085718.

5. A. Geevarghese, G. Wollstein, H. Ishikawa, J. S. Schuman, Optical Coherence Tomography and Glaucoma. Annu. Rev. Vis. Sci. 7 (1), 693–726 (2021), doi: 10.1146/annurev-vision-100419-111350, https://www.annualreviews.org/doi/10.1146/annurev-vision-100419-111350.

6. D. Huang, et al., Optical Coherence Tomography (2015).

7. S. Frenkel, J. E. Morgan, E. Z. Blumenthal, Histological measurement of retinal nerve fibre layer thickness. Eye 19 (5), 491–498 (2005), doi:10.1038/sj.eye.6701569, https://www.nature.com/articles/6701569.

8. J. Liang, D. R. Williams, Aberrations and retinal image quality of the normal human eye. J. Opt. Soc. Am. A 14 (11), 2873 (1997), doi:10.1364/JOSAA.14.002873, https://opg.optica.org/abstract.cfm?URI=josaa-14-11-2873.

9. E. A. Rossi, et al., Imaging individual neurons in the retinal ganglion cell layer of the living eye. Proc. Natl. Acad. Sci. U.S.A. 114 (3), 586–591 (2017), doi:10.1073/pnas.1613445114, https://pnas.org/doi/full/10.1073/pnas.1613445114.

10. Z. Liu, K. Kurokawa, F. Zhang, J. J. Lee, D. T. Miller, Imaging and quantifying ganglion cells and other transparent neurons in the living human retina. Proceedings of the National Academy of Sciences 114 (48), 12803–12808 (2017), doi:10.1073/pnas.1711734114, https://www.pnas.org/doi/full/10.1073/pnas.1711734114.

11. P. Mecê, J. Scholler, K. Groux, et al., Adaptive glasses wavefront sensorless full-field OCT for high-resolution retinal imaging over a wide field-of-view, in Ophthalmic Technologies XXXI (SPIE), vol. 11623 (2021), p. 116230R, doi:10.1117/12.2577969.

12. Y. Cai, O. Thouvenin, K. Grieve, P. Mecê, Influence of static and dynamic ocular aberrations on full-field optical coherence tomography for in vivo high-resolution retinal imaging. Optics Letters 49 (8), 2209–2212 (2024), doi:10.1364/OL.521930.

13. S. K. Gardiner, S. Demirel, J. Reynaud, B. Fortune, Changes in Retinal Nerve Fiber Layer Reflectance Intensity as a Predictor of Functional Progression in Glaucoma. Invest Ophthalmol Vis Sci 57 (3), 1221–1227 (2016), doi:10.1167/iovs.15-18788, https://pmc.ncbi.nlm.nih.gov/articles/PMC4794083/.

14. W. H. Swanson, B. J. King, S. A. Burns, Interpreting Retinal Nerve Fiber Layer Reflectance Defects Based on Presence of Retinal Nerve Fiber Bundles. Optom Vis Sci 98 (5), 531–541 (2021), doi:10.1097/OPX.0000000000001690, https://pmc.ncbi.nlm.nih.gov/articles/PMC8132612/.

15. C. K. S. Leung, et al., Diagnostic assessment of glaucoma and non-glaucomatous optic neuropathies via optical texture analysis of the retinal nerve fibre layer. Nat. Biomed.Eng 6 (5), 593–604 (2022), doi:10.1038/s41551-021-00813-x, https://www.nature.com/articles/s41551-021-00813-x.

16. N. Jansonius, J. Schiefer, J. Nevalainen, J. Paetzold, U. Schiefer, A mathematical model for describing the retinal nerve fiber bundle trajectories in the human eye: Average course, variability, and influence of refraction, optic disc size and optic disc position. Experimental eye research 105 (2012), doi:10.1016/j.exer.2012.10.008.

17. F. Schwarzhans, et al., Automatic retinal nerve fiber bundle tracing based on large field of view polarization sensitive OCT data. Biomed. Opt. Express 13 (1), 65 (2022), doi:10.1364/BOE.443958, https://opg.optica.org/abstract.cfm?URI=boe-13-1-65.

18. R. R. Parakkel, et al., Retinal Nerve Fiber Layer Damage Assessment in Glaucomatous Eyes Using Retinal Retardance Measured by Polarization-Sensitive Optical Coherence Tomography. Trans. Vis. Sci. Tech. 13 (5), 9 (2024), doi:10.1167/tvst.13.5.9, https://tvst.arvojournals.org/article.aspx?articleid=2793660.

19. R. Caro, et al., In vivo imaging of mitochondrial function in normal, glaucoma suspect, and glaucoma eyes. PLoS ONE 20 (1), e0317354.(2025), doi:10.1371/journal.pone.0317354, https://dx.plos.org/10.1371/journal.pone.0317354.

20. R. S. Harwerth, L. Carter-Dawson, F. Shen, E. L. Smith III, M. L. J. Crawford, Ganglion Cell Losses Underlying Visual Field Defects from Experimental Glaucoma. Invest. Ophthalmol. Vis. Sci. 40 (10), 2242–2250 (1999).

21. H. Jayaram, M. Kolko, D. S. Friedman, G. Gazzard, Glaucoma: now and beyond. The Lancet 402 (10414), 1788–1801 (2023), doi:10.1016/S0140-6736(23)01289-8, https://www.sciencedirect.com/science/article/pii/S0140673623012898.

22. C. S. Langlo, A. Amin, S. S. Park, Optical coherence tomography retinal imaging: narrative review of technological advancements and clinical applications. Ann Transl Med 13 (2), 17–17 (2025), doi:10.21037/atm-24-211, https://atm.amegroups.com/article/view/137646/html.

23. R. S. Jonnal, et al., A Review of Adaptive Optics Optical Coherence Tomography: Technical Advances, Scientific Applications, and the Future. Invest Ophthalmol Vis Sci 57 (9), OCT51–OCT68 (2016), doi:10.1167/iovs.16-19103, https://pmc.ncbi.nlm.nih.gov/articles/PMC4968917/.

24. Q.-M. Chi, et al., Evaluation of the Effect of Aging on the Retinal Nerve Fiber Layer Thickness Using Scanning Laser Polarimetry. Journal of Glaucoma 4 (6),406 (1995), https://journals.lww.com/glaucomajournal/abstract/1995/12000/Evaluation_of_the_Effect_of_Aging_on_the_Retinal.6.aspx?casa_token=MSfz6zWS244AAAAA:NW4bRER5v0Jd3sWuV3Co2f09lOkt5NhWZB1klt6-VCS_ronK-RCwYon5fIhn9uJgiFeU925fJImNBoen0W8S75v7Gw.

25. D. Poinoosawmy, L. Fontana, J. X. Wu, F. W. Fitzke, R. A. Hitchings, Variation of nerve fibre layer thickness measurements with age and ethnicity by scanning laser polarimetry. British Journal of Ophthalmology 81 (5), 350–354 (1997), doi:10.1136/bjo.81.5.350, https://bjo.bmj.com/content/81/5/350.

26. C. Bowd, et al., Imaging of the optic disc and retinal nerve fiber layer: the effects of age, optic disc area, refractive error, and gender. J. Opt. Soc. Am. A 19 (1), 197 (2002), doi:10.1364/JOSAA.19.000197, https://opg.optica.org/abstract.cfm?URI=josaa-19-1-197.

27. I. Pereira, et al., Multivariate Model of the Intersubject Variability of the Retinal Nerve Fiber Layer Thickness in Healthy Subjects. Invest. Ophthalmol. Vis. Sci. 56 (9), 5290 (2015), doi:10.1167/iovs.15-17346, http://iovs.arvojournals.org/article.aspx?doi=10.1167/iovs.15-17346.

28. R. S. Parikh, et al., Normal Age-Related Decay of Retinal Nerve Fiber Layer Thickness. Ophthalmology 114 (5), 921–926 (2007), doi:10.1016/j.ophtha.2007.01.023, https://linkinghub.elsevier.com/retrieve/pii/S0161642007001170.

29. J. Scholler, K. Groux, K. Grieve, C. Boccara, P. Mecê, Adaptive-glasses time-domain FFOCT for wide-field high-resolution retinal imaging with increased SNR. Optics Letters 45 (21), 5901–5904 (2020), doi:10.1364/OL.403135.

30. L. Wang, J. Dong, G. Cull, B. Fortune, G. A. Cioffi, Varicosities of Intraretinal Ganglion Cell Axons in Human and Nonhuman Primates. Invest. Ophthalmol. Vis. Sci. 44 (1), 2–9 (2003), doi:10.1167/iovs.02-0333, http://doi.org/10.1167/iovs.02-0333.

31. G. Huang, T. J. Gast, S. A. Burns, In Vivo Adaptive Optics Imaging of the Temporal Raphe and Its Relationship to the Optic Disc and Fovea in the Human Retina. Invest. Ophthalmol. Vis. Sci. 55 (9), 5952–5961 (2014), doi:10.1167/iovs.14-14893, http://doi.org/10.1167/iovs.14-14893.

32. O. P. Kocaoglu, et al., Imaging retinal nerve fiber bundles using optical coherence tomography with adaptive optics. Vision Res 51 (16), 1835–1844 (2011), doi:10.1016/j.visres.2011.06.013, https://pmc.ncbi.nlm.nih.gov/articles/PMC3191496/.

33. Y. Zhang, A. Roorda, Evaluating the lateral resolution of the adaptive optics scanning laser ophthalmoscope. JBO 11 (1), 014002 (2006), doi:10.1117/1.2166434, https://www.spiedigitallibrary.org/journals/journal-of-biomedical-optics/volume-11/issue-1/014002/Evaluating-the-lateral-resolution-of-the-adaptive-optics-scanning-laser/10.1117/1.2166434.full.

34. B. S. Ashimatey, B. J. King, V. E. Malinovsky, W. H. Swanson, Novel Technique for Quantifying Retinal Nerve Fiber Bundle Abnormality in the Temporal Raphe. Optometry and Vision Science 95 (4), 309–317 (2018), doi:10.1097/OPX.0000000000001202.

35. P. Mecê, et al., Coherence gate shaping for wide field high-resolution in vivo retinal imaging with full-field OCT. Biomedical Optics Express 11 (9), 4928–4941 (2020), doi:10.1364/BOE.397066.

36. Y. Cai, K. Grieve, P. Mecê, Characterization and Analysis of Retinal Axial Motion at High Spatiotemporal Resolution and Its Implication for Real-Time Correction in Human Retinal Imaging. Frontiers in Medicine 9 (2022), doi:10.3389/fmed.2022.868217.

37. K. Takayama, et al., High-Resolution Imaging of the Retinal Nerve Fiber Layer in Normal Eyes Using Adaptive Optics Scanning Laser Ophthalmoscopy. PLoS ONE 7 (3), e33158 (2012), doi:10.1371/journal.pone.0033158, https://dx.plos.org/10.1371/journal.pone.0033158.

38. W. H. Swanson, B. J. King, S. A. Burns, Within-subject variability in human retinal nerve fiber bundle width. PLoS ONE 14 (10), e0223350 (2019), doi:10.1371/journal.pone.0223350, https://dx.plos.org/10.1371/journal.pone.0223350.

39. C. Guan, et al., Quantitative Microstructural Analysis of Cellular and Tissue Remodeling in Human Glaucoma Optic Nerve Head. Invest. Ophthalmol. Vis. Sci. 63 (11), 18 (2022), doi:10.1167/iovs.63.11.18, https://doi.org/10.1167/iovs.63.11.18.

40. M. Schneider, R. Fuchshofer, The role of astrocytes in optic nerve head fibrosis in glaucoma. Experimental Eye Research 142, 49–55 (2016), doi:10.1016/j.exer.2015.08.014, https://www.sciencedirect.com/science/article/pii/S0014483515300105.

41. R. F. Spaide, et al., Lateral Resolution of a Commercial Optical Coherence Tomography Instrument. Transl Vis Sci Technol 11 (1), 28 (2022), doi:10.1167/tvst.11.1.28, https://pmc.ncbi.nlm.nih.gov/articles/PMC8787587/.

42. A. Dubois, L. Vabre, A.-C. Boccara, E. Beaurepaire, High-resolution full-field optical co-erence tomography with a Linnik microscope. Appl Opt 41 (4), 805–812 (2002), doi: 10.1364/ao.41.000805.

43. A. Dubois, C. Boccara, [Full-field OCT]. Med Sci (Paris) 22 (10), 859–864 (2006), doi: 10.1051/medsci/20062210859.

44. Y. Rui, et al., Label-Free Imaging of Inflammation at the Level of Single Cells in the Living Human Eye. Ophthalmology Science 4 (5), 100475 (2024), doi:10.1016/j.xops.2024.100475, https://linkinghub.elsevier.com/retrieve/pii/S2666914524000113.

45. D. M. W. Lee, M. Zhang, V. C. Snyder, E. A. Rossi, Multi-spectral autofluorescence variability of the individual retinal pigmented epithelial cells in healthy aging eyes. Sci Rep 14 (1), 30012 (2024), doi:10.1038/s41598-024-81433-8, https://www.nature.com/articles/s41598-024-81433-8.

46. W.-D. Zhou, et al., Cone Mosaic in Eyes with Varied Axial Length Using Adaptive Optics Scanning Laser Ophthalmoscopy (2024), doi:10.21203/rs.3.rs-5471967/v1, https://www.researchsquare.com/article/rs-5471967/v1, iSSN: 2693-5015.

47. Y. Wu, et al., Case Report: novel GUCA1B and ABHD12 mutations in retinitis pigmentosa sine pigmento: expanding the genotypic spectrum through multimodal phenotyping. Front. Med. 12 (2025), doi:10.3389/fmed.2025.1622343, https://www.frontiersin.org/journals/medicine/articles/10.3389/fmed.2025.1622343/full.

48. J. Schindelin, et al., Fiji: an open-source platform for biological-image analysis. Nat Methods 9 (7), 676–682 (2012), doi:10.1038/nmeth.2019, https://www.nature.com/articles/nmeth.2019.

49. P. Mecê, J. Scholler, K. Groux, C. Boccara, High-resolution in-vivo human retinal imaging using full-field OCT with optical stabilization of axial motion. Biomed. Opt. Express, BOE 11 (1), 492–504 (2020), doi:10.1364/BOE.381398, https://opg.optica.org/boe/abstract.cfm?uri=boe-11-1-492.

50. P. Mecê, et al., Real-time optical stabilization of retinal motion at micrometer precision using Adaptive Optics Flood-Illumination Ophthalmoscope (2024), https://hal.science/hal-04742069,hALpreprint,hal-04742069.

51. M. Zhang, et al., Strip-based digital image registration for distortion minimization and robust eye motion measurement from scanned ophthalmic imaging systems. Biomed. Opt. Express, BOE 12 (4), 2353–2372 (2021), doi:10.1364/BOE.418070, https://opg.optica.org/boe/abstract.cfm?uri=boe-12-4-2353.

52. K. V. Vienola, et al., Microstructure of the retinal pigment epithelium near-infrared autofluorescence in healthy young eyes and in patients with AMD. Sci Rep 10 (1), 9561 (2020), doi:10.1038/s41598-020-66581-x, https://www.nature.com/articles/s41598-020-66581-x.

53. Q. Yang, et al., Calibration-free sinusoidal rectification and uniform retinal irradiance in scanning light ophthalmoscopy. Opt. Lett. 40 (1), 85 (2015), doi:10.1364/OL.40.000085, https://opg.optica.org/abstract.cfm?URI=ol-40-1-85.

54. A. F. Frangi, W. J. Niessen, K. L. Vincken, M. A. Viergever, Multiscale vessel enhancement filtering pp. 130–137 (1998), doi:10.1007/BFb0056195.

55. J. Burke, et al., SLOctolyzer: Fully Automatic Analysis Toolkit for Segmentation and Feature Extracting in Scanning Laser Ophthalmoscopy Images. Trans. Vis. Sci. Tech. 13 (11), 7 (2024), https://github.com/jaburke166/SLOctolyzer, doi:10.1167/tvst.13.11.7, https://tvst.arvojournals.org/article.aspx?articleid=2802220.

