## Supplementary text for "High Resolution Multi-depth Quantification of the Retinal Nerve Fiber Layer"

Clémentine Callet* *et al.*,

**This PDF file includes:**

Supplementary Text

### Supplementary Text

### Pre-Processing and exclusion of noisy images for FFOCT only

Pre-Processing starts with a classical 2-phase demodulation scheme to go from the direct images to tomographic images. The stack is then divided per depth offset, giving 11 sub-stacks. For each sub-stack, noisy and poor quality images were detected using rudimentary intensity-based thresholds th1 and th2: $th_1 = 1.02 \times \min_{i \in [1,...,m]} \left( \text{mean}(\text{image}[i]) \right), th_2 = 900$ $i$ the index of the image and $m$ the number of images in the sub-stack. The value of th2 was found empirically by an overall histogram analysis. It excludes images which contain bright spots of dust due to a poor demodulation result caused by large movement of the retina or tracking errors. As a result, images selected verify : $I = \{ i \in [1, n] \mid th_1 < \text{image}(i) < th_2 \}$. All the indicies of resulting good images are saved in a list.

### Reference frame selection by clustering for FFOCT only

Reference frame selection was performed using cross correlation based clustering. Each stack was first divided into groups of 20 consecutive images. For each group only the valid indices corresponding to reliable images were used. When at least one valid image was available in a group, an average image was computed. Otherwise, the averaged image is set to zero. We then characterized the intensity of each averaged image within its central region, excluding the 200 outermost pixels along each border. The resulting mean signal vector is then used to keep the index of the brightest frame from every set of 8 successive averaged images. Each selected index defines a cluster. The total number of clusters obtained is then $n_{\text{cluster}} = n_{\text{groups}}/8 = 800/20/8 = 5$. This ensures clusters are among the selected images based on their mean intensity.

To avoid redundancy, a phase correlation approach was applied between the clusters to estimate their relative displacements. This method provides, for each cluster pair, a displacement vector and an associated correlation coefficient. From these quantities, two metrics were derived: the Euclidean distance of displacement and a binary map identifying reliable correlations. For each image, a map is defined and binary thresholding is applied according to the following conditions: the value of the map at position x and y is 1 when the displacement metric is less than or equal to two and the correlation coefficient is greater than 0.1; otherwise, it is 0. This step allows for the identification of pairs of clusters whose displacement is small and correlation sufficiently high to be considered as valid. The selection proceeds from the last cluster to the first and searches for the set of valid connections in the binary map. If at least one valid connection exists, the corresponding image index is selected as a reference. All connections to this cluster are then removed from the map to avoid redundancy. If no valid connection is found the corresponding reference value is set to zero. Such a clustering allows us to find from 1 to 5 clusters representing a different layer per step. At the end of a full processing of 8000 images we obtain 11 to 55 clusters depending on the volunteer eye motion and tracking efficiency which is driven by SDOCT signal. This clustering ensures that good representative reference images are identified based on correlation and displacements.
